# Pathogenic CD8 T cells defined by longitudinal liver sampling in chronic hepatitis B patients starting antiviral therapy

**DOI:** 10.1101/2021.12.16.21267870

**Authors:** Shirin Nkongolo, Deeqa Mahamed, Adrian Kuipery, Juan D. Sanchez Vasquez, Samuel C. Kim, Aman Mehrotra, Anjali Patel, Christine Hu, Ian McGilvray, Jordan J. Feld, Scott Fung, Diana Chen, Jeffrey J. Wallin, Anuj Gaggar, Harry L. A. Janssen, Adam J. Gehring

**Affiliations:** Toronto Centre for Liver Disease, Toronto General Hospital Research Institute, University Health Network, ON, Canada; Department of Immunology, University of Toronto, ON, Canada; Gilead Sciences, Foster City, CA, USA; Multi-Organ Transplant Program, Toronto General Hospital Research Institute, Toronto, ON, Canada

**Keywords:** Hepatitis B virus (HBV), tenofovir alafenamide (TAF), intrahepatic, inflammation, tissue damage, pathogenesis, Fas ligand (FasL), fine-needle aspiration biopsy, single-cell RNA sequencing, CXCR6, HepG2-NTCP

## Abstract

Accumulation of activated immune cells results in non-specific hepatocyte killing in chronic hepatitis B (CHB), leading to fibrosis and cirrhosis. We enrolled 15 CHB patients with active liver damage to receive antiviral therapy, and performed longitudinal liver sampling using fine-needle aspiration to investigate mechanisms of CHB pathogenesis in the human liver. Single-cell sequencing of total liver cells revealed a distinct liver-resident, polyclonal CD8 T cell population that was enriched at baseline and displayed a highly activated immune signature during liver damage. Cytokine combinations, identified by *in silico* prediction of ligand-receptor interaction, induced the activated phenotype in healthy liver CD8 T cells, resulting in non-specific Fas ligand-mediated killing of target cells. These results define a CD8 T cell population in the human liver that can drive pathogenesis, and a key pathway involved in their function in CHB patients.

## Introduction

Non-specific activation of memory CD8 T cells is a common feature of human viral diseases, such as acute infection with Epstein-Barr virus (EBV) and influenza virus, and chronic infection with human immunodeficiency virus-1 (HIV-1), hepatitis C virus (HCV), hepatitis B virus (HBV), and hepatitis D virus (HDV) (Odumade et al., 2012; Sckisel et al., 2014; Bastidas et al., 2014; Younes et al., 2016; Alanio et al., 2015; Maini et al., 2000; Kefalakes et al., 2021). Bystander-activated CD8 T cells respond in a T cell receptor (TCR)-independent manner and can contribute to protective immunity, e.g. by secretion of interferon-γ (IFNγ); or mediate host tissue damage through cytotoxic mechanisms (Kim and Shin, 2019; Lee et al., 2021). Tissue damage mediated by non-specific activation of CD8 T cells is not restricted to viral infections and has also been reported in type 1 diabetes (Zhang et al., 2010), graft-versus-host disease (Karimi et al., 2015), celiac disease (Meresse et al., 2004), and Lyme arthritis (Whiteside et al., 2018). Therefore, despite their critical role in host protection, dysregulation of CD8 T cell activation can drive immune-mediated pathogenesis, particularly in chronic diseases.

Persistent liver damage is the primary driver of disease progression in chronic HBV infection. Non-specific liver injury ultimately leads to fibrosis, cirrhosis and hepatocellular carcinoma, with an estimated 820,000 HBV-related deaths per year (Chen and Yang, 2011; Fattovich et al., 2004; Razavi-Shearer et al., 2018; World Health Organization, 2021). The kinetics and mechanisms of liver damage have been delineated in animal models of acute HBV infection. Functional HBV-specific T cells produce IFNγ, which induces the chemokines CXCL9 and CXCL10 to coordinate recruitment of inflammatory immune cells (Isogawa et al., 2005; Kakimi et al., 2001). CD8 T cells are the primary mediators of non-specific hepatocyte killing in HBV-infected chimpanzees, and Fas ligand (FasL) was identified as a key non-specific cytotoxic effector molecule in a mouse hydrodynamic HBV infection model (Thimme et al., 2003; Yang et al., 2010).

In contrast to acute infection models, HBV-specific CD8 T cells are deleted in chronic hepatitis B (CHB) patients due to persistent antigen exposure and found at very low frequencies. The remaining HBV-specific CD8 T cells from CHB patients display an exhausted phenotype, arguing against their potential to directly kill infected hepatocytes, or to trigger the inflammatory cascade observed in animal models (Boni et al., 2007; Hoogeveen et al., 2019; Gehring and Protzer, 2019). Therefore, the role of CD8 T cells in non-specific liver damage, the source of IFNγ to initiate the inflammatory infiltrate, and the mechanisms responsible for CD8 T cell-mediated liver damage during chronic HBV infection in the human liver remain poorly defined (Maini et al., 2000; Lee et al., 2021).

A major obstacle to understanding CHB pathogenesis is the virus’ narrow host range and lack of chronic infection models that mimic life-long infection to study pathogenic mechanisms at the site of infection. As such, available animal models with the necessary immunological tools cannot be infected with human HBV and/or do not faithfully recapitulate the pathogenic processes (Dandri and Petersen, 2017; Hu et al., 2019). To overcome these obstacles, we used liver fine-needle aspiration (FNA) (Gill et al., 2018; Gill et al., 2019) to longitudinally sample the livers of CHB patients with active liver damage who started antiviral therapy with the nucleotide analogue tenofovir alafenamide (TAF). TAF treatment inhibits viral replication and reduces liver damage, allowing us to quantify dynamic changes in the intrahepatic immunological landscape at the individual cell level using single-cell RNA sequencing (scRNAseq). This approach identified a highly activated, tissue-resident CD8 T population that causes FasL-dependent apoptosis in hepatoma cells, consistent with their role in liver damage, and expresses IFNγ, which can drive the inflammatory infiltrate. This indicates that innate inflammatory cytokines can drive bystander CD8 T cell activation to initiate the inflammatory process in CHB patients without a requirement for IFNγ derived from exhausted HBV-specific T cells. Defining the CD8 T population mediating liver damage contributes to the understanding of human liver immunopathogenesis and may provide a specific target population to improve antiviral responses (IFNγ) or manage liver damage (FasL).

## Results

### Collection of longitudinal liver FNAs corresponding to response to antiviral therapy

To longitudinally study changes in intrahepatic immune activation associated with liver damage, we enrolled 15 patients with active CHB with elevated alanine aminotransferase (ALT) levels, mean ALT of 4.4 × upper limit of normal (ULN; range: 1.1– 21.8 × ULN), to receive 25 mg/d TAF (figure 1A). TAF is a nucleotide analogue that inhibits HBV replication and halts liver damage but does not eliminate the viral covalently closed circular DNA (cccDNA). Baseline characteristics are summarized in suppl. Table 1. ALT levels decreased by a mean of 3.5-fold (up to 20-fold) by week 12 and by a mean of 5.6-fold (up to 21-fold) by week 24 (figure 1B). At week 24, 12 patients had normalized ALT, with all patients ranging between 0.4–1.3 × ULN. HBV DNA levels ranged between 1.89 × 10^4^–7.97 × 10^7^ IU/ml (mean: 4.84 × 10^6^ IU/ml) at screening and decreased below detection limit in 7 patients, with a maximum of 285 IU/ml and a mean of 14 IU/ml in all patients. As expected, HBsAg levels did not significantly decrease. Out of 7 HBeAg-positive patients at baseline, one underwent HBeAg seroconversion by week 12.

**Figure 1.**
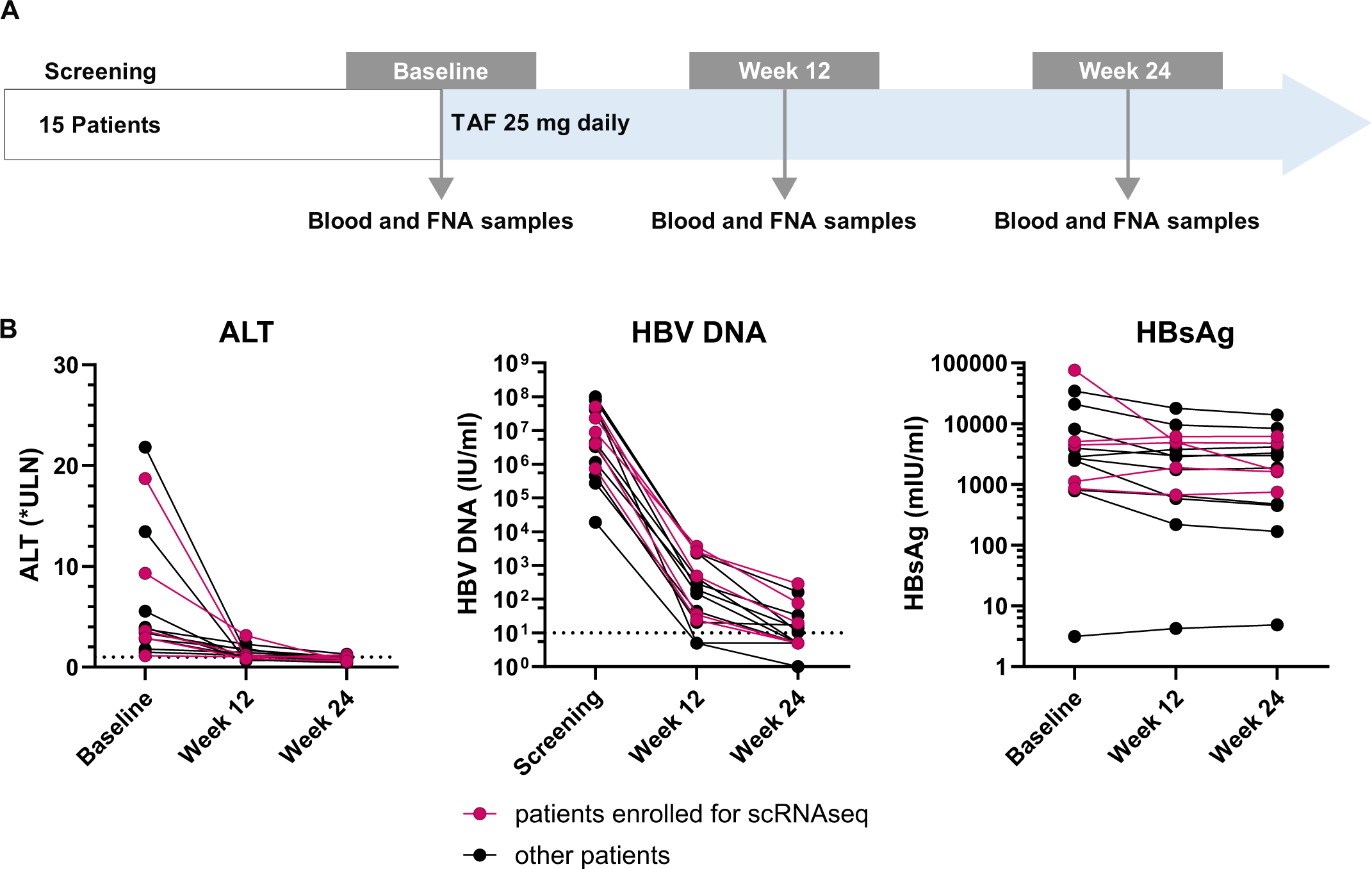
Study design and blood markers of HBV infection under tenofovir alafenamide (TAF) therapy. (A) 15 chronic hepatitis B (CHB) patients with elevated alanine aminotransferase (ALT) levels started nucleoside analogue therapy with TAF 25 mg/d. At baseline and after 12 and 24 weeks of therapy, blood and liver fine-needle aspirates (FNAs) were collected. Longitudinal FNAs from 5 patients were subjected to single-cell RNA sequencing (scRNAseq). (B) ALT, HBV DNA and HBsAg levels in the blood over time. Patients indicated in red are those whose samples were analyzed by scRNAseq.

### ScRNAseq of liver FNAs captures a comprehensive immunological picture of the human liver

Thus far, liver FNAs have only been evaluated using flow cytometry to identify immune cell types based on conventional markers (Spaan et al., 2015; Pembroke et al., 2015; Gill et al., 2019; Yuksel et al., 2021). Therefore, the full cellular diversity of liver FNAs has not been mapped and, given the fragility of some cell types (hepatocytes) or adhesion to the parenchyma (macrophages, endothelial cells, stellate cells), it was not obvious if we could capture cellular diversity in serially collected FNAs to study changes in activation status by scRNAseq. ScRNAseq data from longitudinal samples for 5 patients before, 12, and 24 weeks after starting TAF therapy were integrated following a pipeline to minimize batch effects and eliminate low-quality cells damaged during processing. In total, 38,036 high-quality cells from all patients at all timepoints were included in the analysis and clustered into 32 distinct populations (figure 2A). Importantly, cells from all clusters showed roughly even distribution among the different patients (suppl. figure 1). These data demonstrate that we can consistently capture diverse immune cell types with longitudinal sampling of the HBV-infected liver using FNAs.

**Figure 2.**
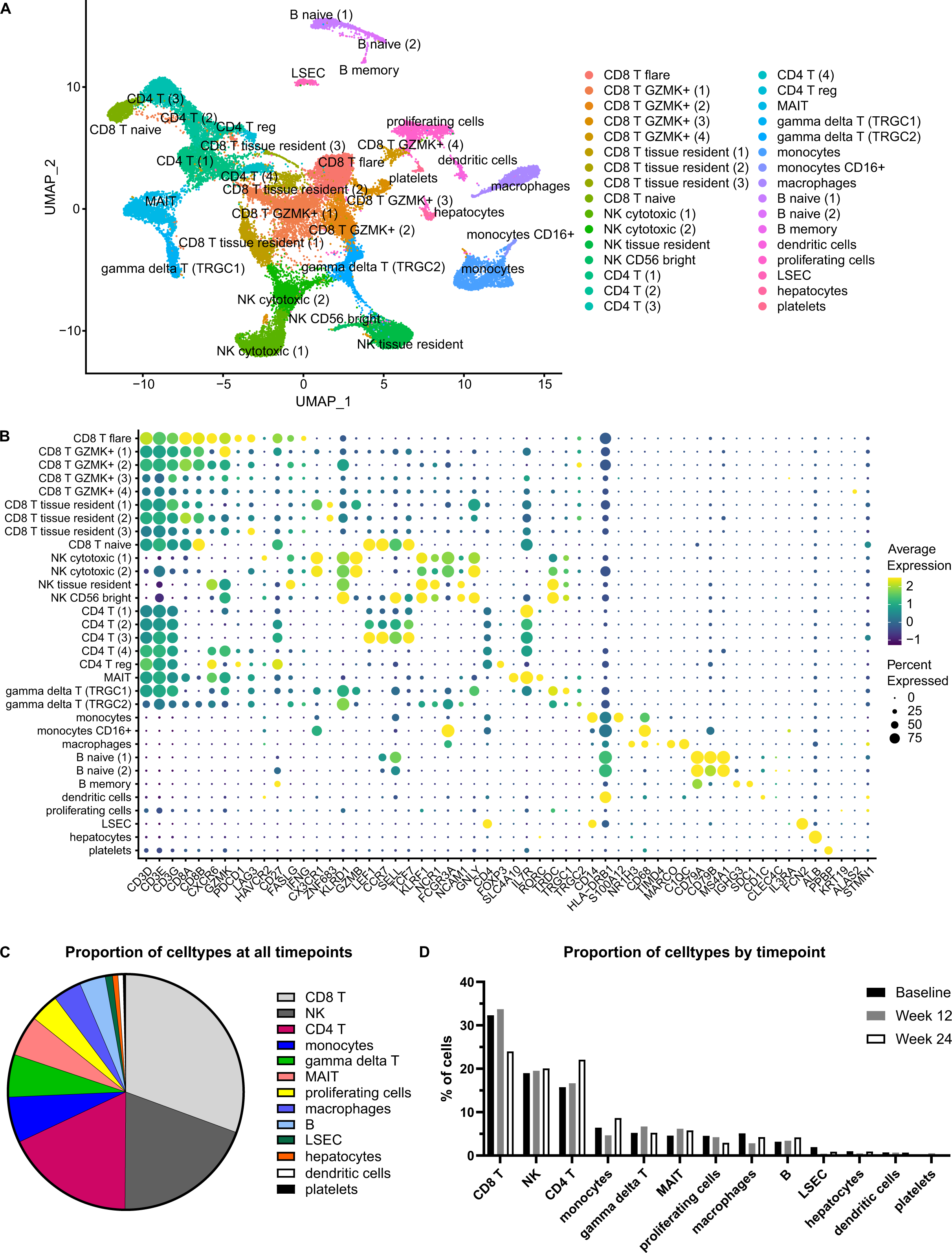
ScRNAseq revealed 32 different populations, with CD8 T cells being the most abundant cell type. (A) Cells from all 5 patients at all 3 timepoints were filtered to exclude low-quality cells and doublets, were clustered, and displayed in a UMAP plot. (B) Cell types were assigned to each cluster using selected marker genes. Whenever the dominant cell type of a cluster was unclear, we used differential gene expression testing and analysis of overexpressed signalling pathways in addition to the displayed marker genes. (C) Distribution of cell types across all timepoints and (D) by timepoint. Note that CD8 T cells are the predominant cell type at all timepoints, but their proportion among all cell types decreases from baseline to week 24.

Cell types were annotated based on differential expression analysis of key lineage and phenotypic marker genes (figure 2B). Cell composition of the liver FNAs was dominated by immune cells (figure 2C). CD8 T cells were most frequent (30.6% of all cells), followed by natural killer (NK) cells (19.5%) and CD4 T cells (17.9%). Less frequent were monocytes (6.3%), γδ T cells (5.9%), mucosal-associated invariant T (MAIT) cells (5.6%), macrophages (3.9%), and B cells (3.6%). The 10x Genomics sequencing approach did not capture significant numbers of granulocytes. Dendritic cells and all non-immune populations – liver sinusoidal endothelial cells (LSEC), hepatocytes, and platelets – each represented less than 1% of all cells. Hepatocytes, which would be expected to be an abundant cell type in liver samples, were largely filtered out as low-quality cells and only accounted for 0.8% of high-quality cells in our dataset, most likely due to limited yield with aspiration alone rather than biopsy, and low stability during sample processing. One cluster of proliferating cells did not show predominant markers for a distinct cell type, but upregulation of cell cycle-associated genes and pathways; this cluster accounted for 4% of all cells.

To determine if the composition of immune cells in the liver changed as TAF therapy suppressed viral load and normalized ALT, we calculated the change in frequency of each population over time. The frequency of CD8 T cells was highest at baseline, during active liver damage, and decreased over time, approaching a significant decline (32.3% of all cells at baseline, compared to 24.0% at week 24; paired t-test: p = 0.06) (figure 2D). By contrast, the proportion of CD4 T cells increased over time (15.7% of all cells at baseline; compared to 22.1% at week 24; paired t-test: p = 0.18). These differences were not statistically significant; but all other cell types showed less change over time, suggesting the enrichment of CD8 T cells at baseline may be physiologically relevant given their known role in liver damage in animal models of HBV pathogenesis.

Overall, we found that FNAs can effectively capture the immunological diversity of the liver across longitudinal samples. While the FNAs did not effectively represent parenchymal cells, such as hepatocytes and endothelial cells, they consistently captured immune cells, allowing us to investigate dynamic changes in transcriptional profiles as ALT levels declined under antiviral treatment.

### Flare CD8 T cells have a unique activated transcriptomic signature

Because CD8 T cells are the primary mediators of HBV-related liver damage in animal models, were the most abundant cell type in our scRNAseq dataset, and were the only cell type that decreased between baseline and week 24, they were analyzed in greater detail. We identified 9 distinct CD8 T populations that differed in their expression of markers related to activation, maturation, trafficking and effector function (figure 3A). One population showed a distinct tissue resident (CXCR6), activated signature including high expression of effector molecules (IFNγ, FasL, granzyme A (GZMA), granzyme K (GZMK), multiple chemokines (CCL3, CCL4, CCL5)), activation markers (4-1BB (CD137/TNFRSF9), CD38, HLA-DR, CD27, CD74), and high expression of exhaustion markers (PD-1 (PDCD1), LAG3, TIGIT), compared to the other 8 clusters. Co-expression analysis for cells that were positive for CXCR6/IFNγ/FasL in this population showed that 76.1% of cells in this cluster expressed CXCR6, 29.9% of cells were IFNγ(+) and 44.0% were FasL(+). Of the cells that were CXCR6(+), 32.2% were IFNγ(+) and 47.1% were FasL(+) (not shown). Given this highly activated phenotype and expression of effector molecules associated with liver damage, we termed this population flare CD8 T cells.

**Figure 3.**
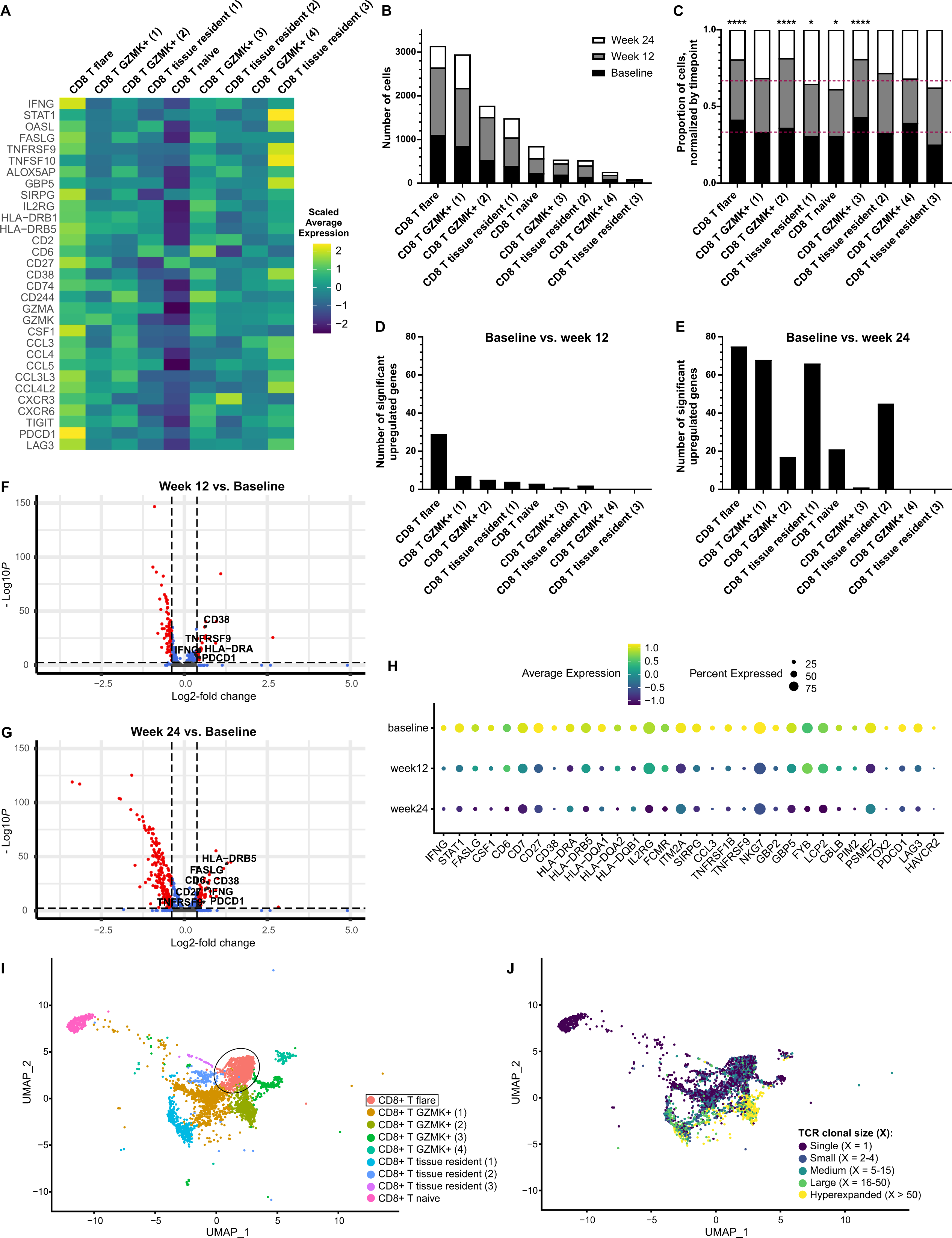
Flare CD8 T cells are a unique inflammatory and polyclonal CD8 T population that down-regulates immune-related genes as ALT levels decline. (A) Heatmap displaying a unique activated immunological signature of flare CD8 T cells at baseline, the time of active liver damage. (B) Numbers of cells in the different CD8 T cell clusters, stratified by timepoint. (C) Proportions of cells in the different CD8 T cell clusters, normalized by the total number of cells at each timepoint. Red lines indicate what would be an equal distribution across all timepoints. Z-test to test for significant enrichment at baseline or week 24: * p < 0.05, **** p < 0.0001. (D) Numbers of differentially upregulated genes in each CD8 T cell cluster, when comparing baseline and week 12, respectively (E) baseline and week 24. (F) Volcano plot showing genes that are differentially expressed over time by 12 weeks, respectively (G) by 24 weeks, in the flare CD8 T population. Thresholds: p < 0.005 and fold change ≥ 1.3. Genes upregulated at baseline are shown to the right side of each plot and heavily overlap between the two comparisons. Genes to the left side of each pot (downregulated at baseline) are mainly ribosomal and mitochondrial genes. (H) Expression of those genes that are differentially upregulated in flare CD8 T cells at baseline, and immune-related, over time. (I) CD8 T cell clusters at the time of active liver damage. UMAP plot, (J) overlaid with the respective clonal sizes of T cell receptor (TCR) clonotypes. For each cell, the dominant TCR sequence was used for analysis. Cells for which no TCR sequencing data was available are shown in (I), but not in (J).

We hypothesized that flare CD8 T cells would be expanded at baseline, during ongoing liver damage, and show the greatest transcriptional changes over time. Therefore, we analyzed the size and relative frequencies of CD8 T cell clusters. Flare CD8 T cells were the largest population and comprised 3,144 cells, representing 27% of all cells in CD8 T cell clusters (figure 3B) (corresponding to 8.3% of all cells, regardless of cell type). This population, along with two other populations (CD8 T GZMK+ (2) and (3)), were significantly enriched in FNA samples taken at baseline (figure 3C). By week 24, when ALT levels had largely normalized, flare CD8 T cells had contracted by 53%. As a measure of changes in transcriptional profiles, we quantified the number of significantly differentially expressed genes (DEGs) that were downregulated from baseline to weeks 12 and 24 after starting therapy. In addition to being enriched at baseline, flare CD8 T cells showed the greatest changes in DEGs as ALT declined. Based on a Bonferroni-corrected adjusted p value < 0.005 and a fold change ≥ 1.3, flare CD8 T cells had 29 genes that were significantly decreased by week 12, when mean ALT had already declined 3.5-fold; whereas all other CD8 T cell clusters had only 0–7 significantly downregulated DEGs between baseline and week 12 (figure 3D). The number of significantly downregulated DEGs in flare CD8 T cells increased to 75 when compared to week 24 (figure 3E). By that time, other CD8 T cell clusters also showed significant downregulation with DEGs ranging between 0-68.

In total, we found 80 unique genes that were significantly upregulated in flare CD8 T cells at baseline, during active liver damage, compared to the later timepoints (figure 3D-G). The largest proportion of these genes was immune-related genes (n = 32), followed by mitochondrial and ribosomal genes (n = 14) and genes regulating transcription/translation (n = 8). By contrast, genes that were upregulated following decline of ALT levels at weeks 12 and 24, compared to baseline, were overwhelmingly mitochondrial and ribosomal genes. The 32 immune-related genes that were significantly upregulated during active liver damage in flare CD8 T cells not only decreased with respect to mean expression levels in the individual cells, but also in the percentages of transcript-positive cells, over time (figure 3H).

Further characterization of the CD8 T cell clusters using TCR sequences of the same cells in the scRNAseq data revealed both polyclonal and hyperexpanded populations (figure 3I-J). Naïve CD8 T cells – as expected – displayed only single clonotypes that were unique for each cell (clonal size of 1). On the other hand, most cells in the CD8 T GZMK+ (2) population were clonally expanded. Flare CD8 T cells were polyclonal, with the vast majority of cells (76%) either having a unique clonotype or a clonotype of clonal size ≤ 4 in each sample, and 93% of cells having a clonotype of clonal size ≤ 15.

Taken together, flare CD8 T cells stood out from all other CD8 T populations due to high expression of activation markers and effector molecules, especially at baseline. This was the largest of all CD8 T populations with relative enrichment at the time of active liver damage, and it displayed a polyclonal TCR composition. It was the only CD8 T population that showed significant downregulation of a substantial number of genes from baseline to week 12, corresponding to the main decrease in ALT levels. Under TAF therapy, the mean expression of immune-related activation genes decreased, including IFNγ and FasL. Therefore, flare CD8 T cells displayed all hallmarks of a CD8 T population with the potential to drive inflammatory events leading to liver damage.

### Validation of the flare CD8 T population at the protein level in liver FNAs

To correlate transcriptomic data to actual protein expression, we developed a multi-color panel designed to identify the flare CD8 T population in cryopreserved longitudinal FNA samples using flow cytometry. Markers and gating strategy are displayed in suppl. figure 2. Due to low cell numbers obtained from the FNAs, we focussed on extracellular markers to minimize cell loss during sample staining. Longitudinal FNAs and matching peripheral blood mononuclear cells (PBMC) from 4 patients were analyzed. As expected, FNA cells contained significantly more tissue resident CXCR6(+) CD8 T cells than PBMC (FNA: mean 29.3% of CD8 T cells were CXCR6(+); PBMC: mean 12.0%; suppl. figure 3A).

Using R packages flowCore and CATALYST, we clustered CD3(+) cells from the different sample types (FNA/PBMC) at baseline or week 24 (figure 4). For FNA samples, stratified by timepoint, a clustering resolution leading to 6 clusters was determined to be optimal since additional clusters did not lead to significant extra cluster stability. In baseline FNAs, three CD8 T populations were identified, as well as two CD4 T populations and one population with low expression of both CD4 and CD8 (figure 4A-B). One CD8 T population had distinctly higher expression of CXCR6, CD38, 4-1BB, FasL and PD-1 than all other clusters. In addition, this population highly expressed CD27 and CD6, overall corresponding well to the flare CD8 T cells defined by scRNAseq. This population could consistently be identified at any resolution between 4–20 clusters. Relative expression of markers for each population is shown in figure 4B. At the resolution of 6 clusters, this flare CD8 T population defined by flow cytometry accounted for 6.5% of all cells in CD8 T cell clusters (corresponding to 2.1% of all CD3(+) cells which were analyzed). By contrast, flare CD8 T cells in scRNAseq data accounted for 27% of all cells in CD8 T cell clusters. However, scRNAseq was done on fresh cells immediately after samples were collected from patients, in contrast to cryopreserved samples analyzed by flow cytometry. Despite the potential impact of cryopreservation on surface marker expression, which can also occur in PBMC samples, we clearly identified a flare CD8 T population in the liver FNA samples at the protein level using flow cytometry.

**Figure 4.**
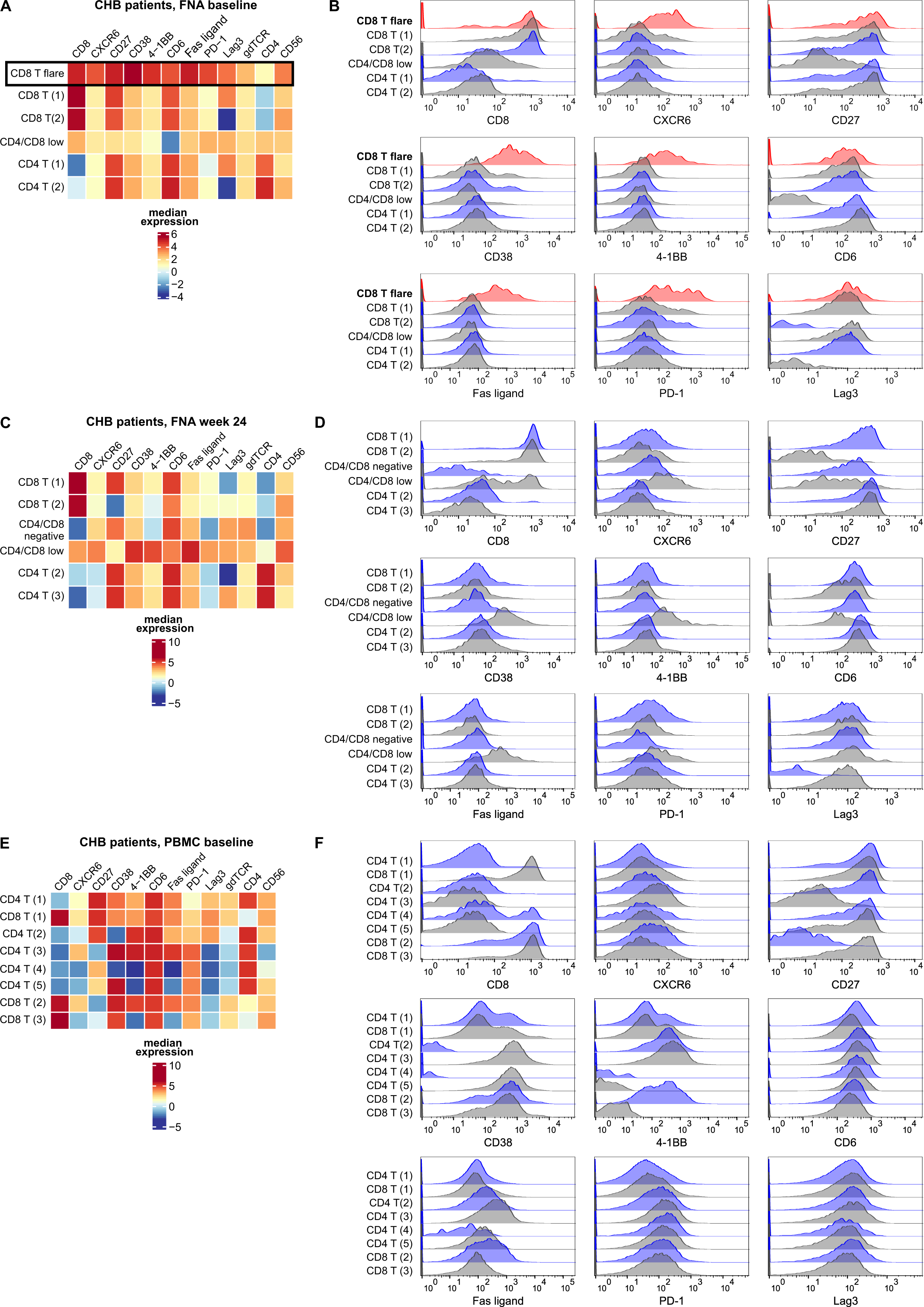
The flare CD8 T population can be identified on the protein level in FNAs from patients with active liver damage, but not after 24 weeks of TAF therapy and not in PBMC. Using a multi-color flow cytometry panel for FNA samples from CHB patients with ongoing liver damage (A-B) and as ALT levels had largely normalized (C-D), and corresponding PBMC samples from CHB patients with active liver damage (E-F), single, live, CD3(+) lymphocytes were selected and clustered. Heatmaps show the median fluorescence intensity; histograms display the distribution of expression levels for CD8 and key flare CD8 T cell phenotypic markers for each cluster.

In contrast to baseline samples, week 24 FNA samples (figure 4C-D) displayed two clusters with high CD8 expression, but low or negative expression of most phenotypic markers defining flare CD8 T cells. One population (CD4/CD8 low) expressed both CD4 and CD8 at low-to-moderate levels, had high expression of CD38, 4-1BB and FasL, but only low-to-moderate levels of the other markers. Expression levels of each marker are displayed in histograms in figure 4D. Consistent with our transcriptional data showing downregulation of immune-related genes at week 24, a clear flare CD8 T cluster could not be identified at the protein level by flow cytometry at week 24. This held true for any clustering resolution between 4-20 clusters.

Lastly, we determined if a similar flare CD8 T population could be identified in matching PBMC from the same patients at baseline. PBMC samples (figure 4E-F) needed to be clustered at a higher resolution to obtain more than one CD8 T cell cluster using the markers contained with the panel. No cluster displayed high expression of CXCR6 or the combination of surface markers that defined flare CD8 T cells (figure 4E). Therefore, a distinct flare CD8 T population could only be identified at the protein level in CHB patient liver FNA samples at the time of active liver damage, but not in liver FNA samples after 24 weeks of TAF therapy, or in matched PBMC samples. This indicates that the flare CD8 T population exists uniquely in the liver during ongoing liver damage.

### IL-2 and IL-12 induce flare CD8 T cells in intrahepatic mononuclear cells (IHMC)

Because flare CD8 T cells were detectable at the protein level only at the time of active liver damage, we anticipated that cytokines present in the inflammatory liver environment were responsible for their activation. Only low numbers of immune cells can be obtained from FNAs: each pass typically yields 20,000–50,000 live cells, with only 4 passes taken from each patient per timepoint. To conduct functional experiments, we therefore took advantage of IHMC harvested from liver perfusion samples. During transplantation, the liver is perfused to remove immune cells from the sinusoids prior to transplant. All IHMC used in this study were collected from living donor liver transplant specimens and were considered healthy without infection or necrosis that could be present in deceased liver donors. The T cell composition of liver perfusions displayed hallmarks expected from liver-derived samples, including an inverted CD4:CD8 ratio (dominant CD8) and increased expression of tissue-associated markers (suppl. figure 3.) In contrast to liver FNAs, liver perfusion samples provide higher cell numbers and originate from non-infected individuals with no significant liver inflammation or liver damage.

We recently defined the broad inflammatory profile that is associated with liver damage in CHB patients (Johnson-Valiente et al., 2021). We hypothesized that flare CD8 T cells engage the inflammatory microenvironment found in the liver of CHB patients to acquire their activated phenotype. Our goal was therefore to define crucial factors to mimic the liver microenvironment and induce a population correlating to flare CD8 T cells in IHMC. We used NicheNet to *in silico* predict factors responsible for upregulation of two key effector genes defining flare CD8 T cells, FasL and IFNγ. In this analysis, signaling molecules, originating from any of the 32 immune cell subsets defined by scRNAseq, were analyzed to predict which of them upregulated FasL and/or IFNγ in flare CD8 T cells at baseline, compared to week 24. Table 1 shows the top 25 ligands with the highest predicted potentials for FasL and IFNγ. Notably, there was a distinct overlap between the predicted ligands for upregulation of both genes.

**Table 1.** Top ligands predicted to upregulate Fas ligand and IFNγ in flare CD8 T cells. NicheNet analysis revealed ligands with the highest regulatory potential score for upregulation at baseline, compared to week 24.

| Ligand | Regulatory potential score for Fas ligand in/on flare CD8 T cells | Ligand | Regulatory potential score for IFNγ in flare CD8 T cells |
| --- | --- | --- | --- |
| IL10 | 0.00765 | IL27 | 0.00863 |
| IL12B | 0.00725 | IL12B | 0.00721 |
| IL12A | 0.00717 | IL12A | 0.00705 |
| TGFB1 | 0.00694 | IL23A | 0.00688 |
| TNF | 0.00664 | EBI3 | 0.00656 |
| IL15 | 0.00646 | IL21 | 0.00559 |
| CD40LG | 0.00545 | IL2 | 0.00528 |
| THBS1 | 0.00532 | IL4 | 0.00514 |
| IL4 | 0.00350 | CD28 | 0.00481 |
| IL27 | 0.00341 | CD86 | 0.00476 |
| IL2 | 0.00339 | IFNA4 | 0.00475 |
| TSLP | 0.00326 | IL1B | 0.00469 |
| IFNL1 | 0.00314 | IFNA7 | 0.00460 |
| IFNG | 0.00311 | IFNA16 | 0.00458 |
| CD28 | 0.00290 | IFNA10 | 0.00455 |
| IL23A | 0.00290 | IFNA6 | 0.00450 |
| ADAM17 | 0.00286 | IFNA14 | 0.00448 |
| CRH | 0.00269 | ICAM1 | 0.00448 |
| CD80 | 0.00264 | TGFB1 | 0.00447 |
| EBI3 | 0.00262 | IFNA2 | 0.00445 |
| EGF | 0.00261 | IFNA8 | 0.00439 |
| KISS1 | 0.00236 | IFNK | 0.00433 |
| LRPAP1 | 0.00236 | IFNA5 | 0.00428 |
| INSL5 | 0.00235 | IFNA17 | 0.00422 |
| IL21 | 0.00219 | IFNA21 | 0.00401 |

In exploratory experiments for *ex vivo* verification, we selected cytokines that had the highest predictive value for upregulation of FasL, but also appeared in the list for IFNγ. We selected IL-2, IL-4, IL-10, IL-12p70 (corresponding to IL-12A and IL-12B), IL-15, IL-21, IL-27, TGFβ, TNF, IFNα and IFNγ (suppl. figure 4A and 4C), and subsequently some combinations thereof (suppl. figure 4B and 4D), to stimulate IHMC for 24 hours. Upregulation of FasL and IFNγ in IHMC-derived CXCR6(+) CD8 T cells was quantified by flow cytometry. The combination of IL-2 + IL-12 was most effective in inducing both proteins.

Among all CXCR6(+) CD8 T cells, the combination of IL-2 and IL-12 treatment increased IFNγ(+) cells from 1.6% to 13.4% and FasL(+) cells from 3.7% to 12.9% and (figure 5A). This can be compared to 19.8% FasL(+) of all CXCR6(+) CD8 T cells by flow cytometry in baseline FNAs from CHB patients (IFNγ as an intracellular cytokine was not assessed by flow cytometry of FNAs due to low cell numbers). We next confirmed that the broader panel of flare CD8 T cell markers was induced by cytokines, using multi-color flow cytometry (gating strategy displayed in suppl. figure 2) on IHMC without and with IL-2 + IL-12 treatment for 24 hours (figure 5B and suppl. figure 5). Untreated IHMC showed 6 CD8 T populations (out of 8 populations in total), none of which had a high expression of all flare CD8 T cell markers. One CD8 T population – CD8 T (5), which accounted for 11.3% of all CD3(+) lymphocytes – expressed flare CD8 T cell markers at a low-to-moderate level. By contrast, after IL-2 + IL-12 treatment, a flare CD8 T population with medium-to-high expression of all markers of interest was identified, which comprised 24.4% of all T cells. These data indicate that exposure to IL-2 + IL-12 can induce a CD8 T population with a phenotype similar to what we observed in liver FNAs from CHB patients during active liver damage.

**Figure 5.**
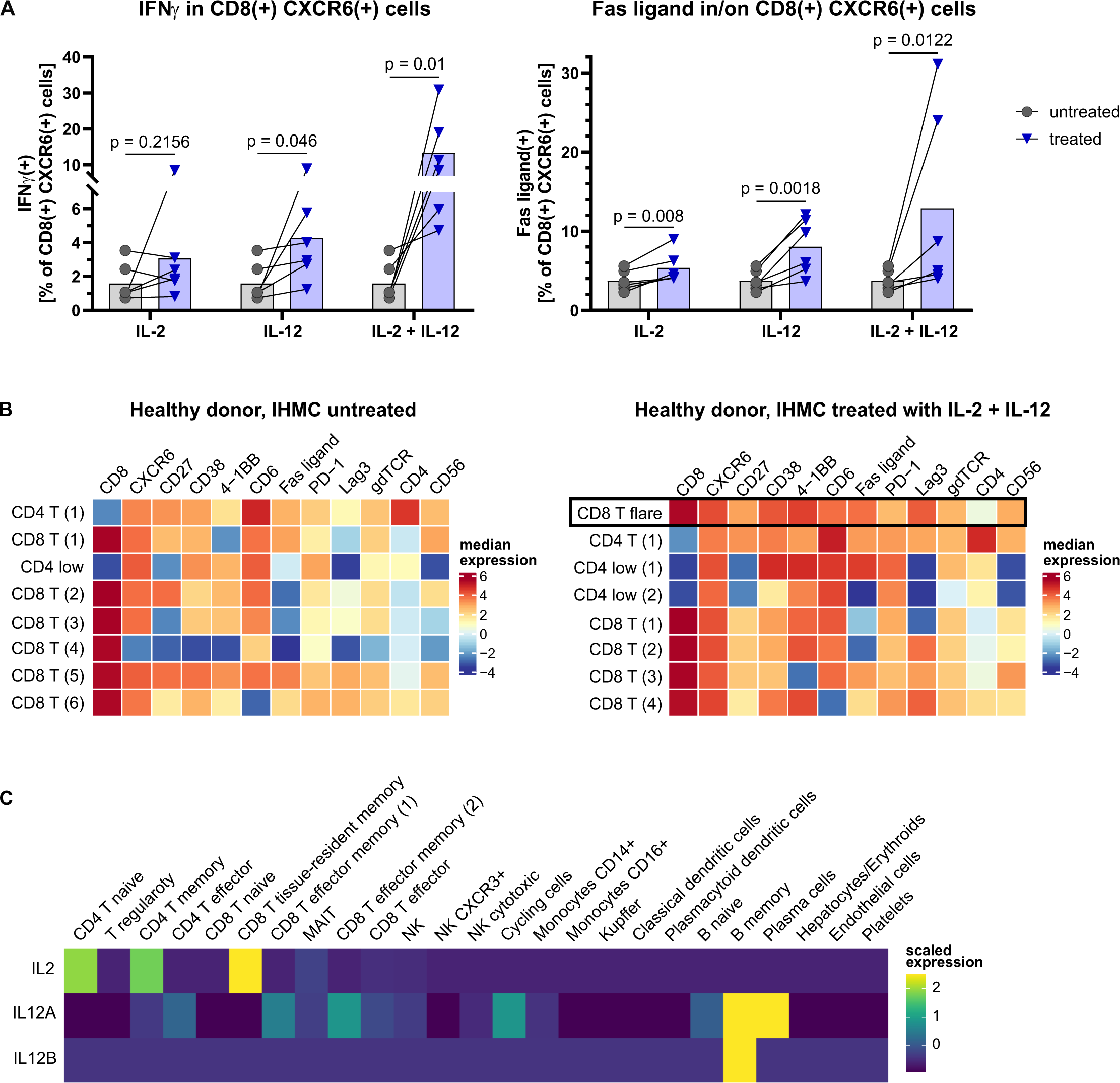
The combination of IL-2 and IL-12 induces flare lCD8 T cells in healthy donor intrahepatic cells. (A) IHMC from 6 healthy donors were treated with IL-2, IL-12, or IL-2 + IL-12 for 24 h before quantification of IFNγ and Fas ligand expression of CXCR6(+) CD8 T cells by flow cytometry. Statistical significance was assessed by paired t test. (B) A multi-color flow cytometry panel was used to analyze IHMC from 6 healthy donors, without and with 24 h treatment with IL-2 + IL-12. Single, live, CD3(+) lymphocytes were selected before clustering. The flare CD8 T population, corresponding to baseline FNA samples from CHB patients, was only found in the treated healthy donor IHMC. Heatmaps show median expression. (C) Source of IL-2 and IL-12 in CHB patients’ livers at baseline. A targeted scRNAseq assay was used to enrich for cytokine genes. Both IL-2 and IL-12A/IL-12B could be detected.

Since IL-2 and IL-12 were sufficient to induce flare CD8 T cells, we hypothesized that these were crucial factors in the CHB patient liver microenvironment during liver damage. There are challenges with detecting cytokines in the 10x Genomics scRNAseq platform. To determine the main source of IL-2 and IL-12 in CHB patients at baseline, we used a targeted gene expression kit for enrichment of 1,056 genes that included all cytokines of interest. We could detect IL-2, IL-12A and IL-12B, both of which are required to form the active form of IL-12. IL-2 was shown to be mainly expressed by CD4 T cells and a tissue resident memory CD8 T population. IL-12 was mainly expressed by B cells (figure 5C). In the scRNAseq data from CHB patients, CD4 T, CD8 T and B cells were therefore identified as likely sources of the cytokines confirmed to induce the flare CD8 T phenotype in healthy IHMC samples.

### IHMC-derived flare CD8 T cells can induce apoptosis in human hepatoma cells in a Fas ligand-dependent manner

We next aimed to determine whether the activation profile defining flare CD8 T cells can induce non-specific liver damage through killing of host hepatocytes, independent of antigen specificity. In the context of non-specific liver damage, FasL binds to Fas on target cells to initiate apoptosis. We used the human hepatoma cell line HepG2-NTCP as a model for hepatocytes. HepG2-NTCP cells were not infected with HBV and not HLA-matched with IHMC donors to investigate non-specific killing independent of HBV antigen presentation (figure 6). CD8 T cells incubated in the absence of cytokines served as a control for background lysis that may occur through allogenic reactivity.

**Figure 6.**
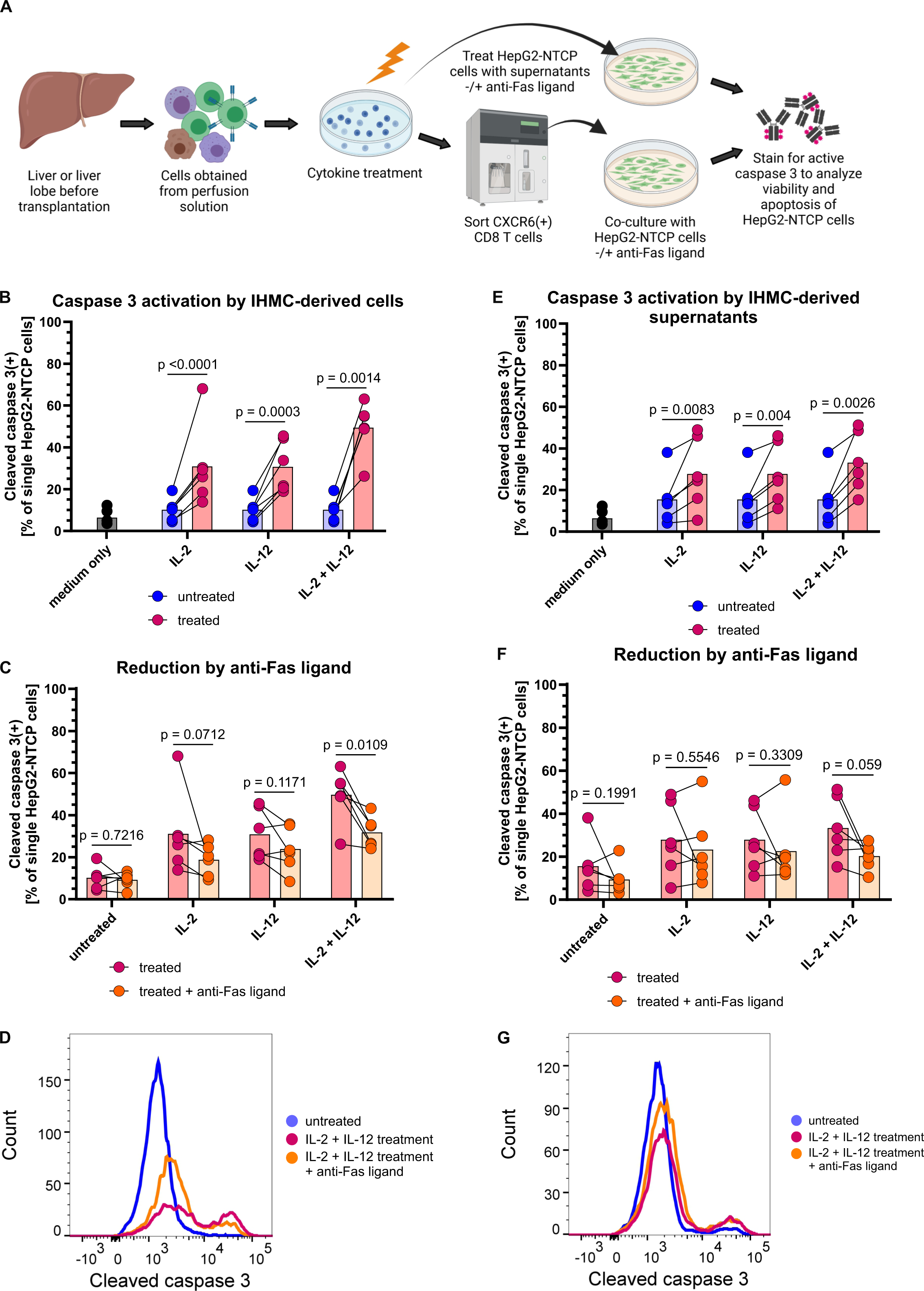
IHMC-derived CD8(+) CXCR6(+) cells corresponding to the flare CD8 T population can induce apoptosis in a hepatoma cell line. (A) Experimental setup: intrahepatic mononuclear cells (IHMCs) from living liver donors were treated with indicated cytokines to induce the population corresponding to flare CD8 T cells. Cells were sorted to obtain the CD8(+) CXCR6(+) subpopulation and co-cultured with HepG2-NTCP cells for 24h. In parallel, HepG2-NTCP cells were cultivated in IHMC-derived supernatants. The potential of cells, respectively supernatants, to induce apoptosis in HepG2-NTCP cells was evaluated by quantifying active caspase 3 using flow cytometry. In addition, we tested whether a neutralizing anti-Fas ligand antibody could inhibit induction of apoptosis. (B) Active caspase 3 in HepG2-NTCP cells that were co-cultured with IHMC-derived CD8(+) CXCR6(+) cells. Dots indicate individual donors; bars indicate mean values. Medium only indicates HepG2-NTCP cells without co-cultivation. (C) Active caspase 3 in HepG2-NTCP cells after co-culture, with and without Fas ligand blockade. (D) Histogram from one representative donor. (E-G) Active caspase 3 in HepG2-NTCP cells that were treated with IHMC-derived supernatants. Ratio paired t test was used to test for statistical significance.

CXCR6 was the one marker that, out of all 24,469 genes in our scRNAseq analysis, correlated best with FasL positivity in each individual flare CD8 T cell (not shown). After cytokine stimulation for 24 h, we sorted CXCR6(+) CD8 T cells to enrich for the CD8 T population containing flare CD8 T cells. We co-cultured the sorted cells with HepG2-NTCP cells for 24 h. In a parallel setting, sorted IHMC-derived CD8(+) CXCR6(+) cells were pre-incubated with a neutralizing anti-FasL antibody before co-cultivation with HepG2-NTCP cells. Caspase 3 cleavage into an active form is a key step in apoptosis pathways. Cleaved caspase 3 was quantified in HepG2-NTCP cells to measure killing. In addition, we used the supernatants derived from IHMC treated with IL-2 and IL-12 for 24 h, without and with prior addition of anti-FasL, to distinguish between contact-dependent killing by flare CD8 T cells and killing through soluble mediators (figure 6A). Six donors were included for individual experiments. HepG2-NTCP cells showed a background of 6.5% active caspase 3 positivity. This proportion increased to 10.2% when hepatoma cells were co-cultured with CD8(+) CXCR6(+) cells derived from untreated IHMC (figure 6B). IL-2 or IL-12 treatment of IHMC led to 31.0% and 30.8%, respectively, of HepG2-NTCP cells with active caspase 3. Notably, when IHMC were treated with IL-2 + IL-12, sorted CD8(+) CXCR6(+) cells induced caspase 3 activation in 49.5% of HepG2-NTCP cells (4.9-fold increase) when measured after 24 h (figure 6B). These data show that CD8 T cells with a flare CD8 T phenotype, activated by IL-2 + IL-12, mediated enhanced killing of HepG2-NTCP cells.

To determine whether flare CD8 T cell-induced hepatoma cell killing was dependent on the Fas/FasL axis, we next tested whether FasL blockade could inhibit the apoptosis-inducing effect (figure 6C-D). There was no significant difference between cells derived from untreated IHMC without or with addition of anti-FasL prior to co-culture. The same held true for cells derived from IHMC treated with either IL-2 or IL-12 alone, although FasL blocking had an effect approaching significance. However, when cells were activated with IL-2 + IL-12, their capacity to kill HepG2-NTCP cells was significantly reduced by blocking FasL (figure 6C). Blocking FasL reduced HepG2-NTCP killing to 31.7%. This means that about half of the apoptosis-inducing effect by IL-2 + IL-12-activated IHMC-derived CXCR6(+) CD8 T cells was abolished through FasL blockade, when adjusted by subtracting killing from untreated cells. Representative histograms for the FasL blocking data are depicted in figure 6D, showing a clear decrease in active caspase 3 staining after blocking FasL. These data show that the Fas/FasL pathway is a key mechanism associated with non-specific, contact-dependent HepG2-NTCP killing by flare CD8 T cells.

To assess if soluble factors induced by the activation of intrahepatic lymphocytes with IL-2 + IL-12 could also mediate cytotoxicity towards HepG2-NTCP cells, we treated HepG2-NTCP cells with supernatants collected from IHMC after 24 h of activation with IL-2 + IL-12. Supernatants had a less pronounced effect on HepG2-NTCP killing but still induced caspase 3 activity (figure 6E-G). The highest effect was found for supernatants from IL-2 + IL-12-treated IHMC, which increased the proportion of active caspase 3(+) cells 2.1-fold. Antagonism with anti-FasL slightly decreased killing, but the decrease was not significant in any of the conditions (figure 6F), and the histogram representation of the data was distinctly different from contact-mediated killing (figure 6D and 6G). This suggests that contact-dependent killing is the primary mechanism but an additional, contact-independent non-specific killing mechanism may also be induced by IL-2 + IL-12 treatment.

In conclusion, IHMC-derived flare CD8 T cells were induced by treatment with IL-2 and IL-12. Co-culture of HepG2-NTCP cells with flare CD8 T cells, and to a lesser extent treatment with IL-2 + IL12-induced IHMC supernatants, substantially activated apoptosis pathways in HepG2-NTCP cells, which could partially be inhibited by antagonizing membrane-bound FasL. These data demonstrate a pathway for contact-dependent killing by flare CD8 T cells, which can promote liver damage in a non-specific manner.

## Discussion

Infiltration of activated CD8 T cells is a hallmark of HBV-associated liver damage in CHB patients (Maini et al., 2000; Traum et al., 2021), but our understanding of their role in viral hepatitis has relied largely on animal models. Longitudinal human liver sampling with core biopsies is impractical and invasive. Therefore, only cross-sectional data have been available to infer immune functionality and pathological mechanisms in humans. Liver FNAs revolutionized our ability to investigate liver-specific mechanisms of immunity and pathogenesis because the minimal risk allows for tissue sampling on time scales consistent with antiviral effects of therapy. We took advantage of liver FNAs to longitudinally quantify dynamic changes in the activation status of individual immune cells in the liver, as initially elevated ALT levels normalized under antiviral therapy. ScRNAseq of liver FNAs provided high-resolution data to define a distinct, activated, liver-resident, polyclonal CD8 T population in CHB patients that was capable of non-specific cytotoxicity through a FasL-dependent pathway. Tracking this population of cells over time suggested that these tissue-resident CXCR6(+) CD8 T cells are the primary mediators of liver damage. Furthermore, our data show that IFNγ, which induces chemokines associated with immune cell infiltration, can be induced in the CXCR6(+) CD8 T cells through non-specific activation. This is an important observation since CHB pathogenesis in all animal models relied on IFNγ produced from fully functional HBV-specific T cells, while HBV-specific CD8 T cells in CHB patients are highly exhausted, and unlikely to drive inflammation. Definition of a specific CD8 T population primarily responsible for liver damage substantially advances the understanding of pathogenic processes in CHB patients.

By conventional understanding, the flare CD8 T cluster displayed a paradoxical phenotype on both transcriptional and protein levels, expressing a broad profile of activation markers, chemokines and cytokines, while also expressing the highest levels of immune checkpoint receptors such as PD-1, LAG-3, TIM-3 and TIGIT. However, PD-1 expression was transient in the scRNAseq data, indicating it being a marker of activation rather than exhaustion. This is consistent with acute HBV infection, where PD-1 positively correlates with ALT levels (Ahn et al., 2018; Peng et al., 2008; Jubel et al., 2020) and non-HBV models of chronic inflammation such as juvenile idiopathic arthritis or polyomavirus encephalitis (Petrelli et al., 2018; Shwetank et al., 2019; Bernard, 2018). Similarly, a recent study found PD-1 upregulation associated with non-specific activation of bystander CD8 T cells in chronic hepatitis D (Kefalakes et al., 2021). These data contrast with the effect of PD-1 expression on HBV-specific CD8 T cells, which, in that context, is a distinct marker of their exhausted functional profile, and negatively correlates with virus-specific CD8 T cell responses (Bertoletti and Gehring, 2006; Boni et al., 2007; Peng et al., 2008). Furthermore, given the size of the flare CD8 T population, and its highly activated profile at baseline, when there was least viral control, it is exceedingly unlikely that these cells represent the HBV-specific T cell response in the CHB patient liver. Taken together, high expression of immune checkpoints such as PD-1 in the flare CD8 T population does not impede CD8 T cell functionality, and expression of these checkpoint inhibitors cannot always be interpreted as CD8 T cell exhaustion in scRNAseq data. Furthermore, our data show that full effector function, including IFNγ production, can be achieved in the presence of PD-1 expression. This has implications for immunotherapy in CHB patients, where modulating the liver environment to overcome immune checkpoints may be more effective, and safer, than systemic administration of checkpoint inhibitor therapy, which can lead to immune-related adverse events.

The ability to mediate clinically significant liver damage, which is reflected by elevated ALT in the serum, requires a sufficient number of CD8 T cells to be activated to kill hepatocytes, and/or the ability of those cells to serially kill multiple hepatocytes. The Fas/FasL system has been associated with liver damage in CHB in mouse models and cell culture of primary human hepatocytes from CHB patients (Galle et al., 1995; Kondo et al., 1997; Kennedy et al., 2001; Brenner et al., 2013). FasL, when membrane-bound, binds to Fas on target cells and induces pro-apoptotic pathways. However, the soluble form may not induce apoptosis, and even protect target cells through competition for the Fas receptor (Schneider et al., 1998; Suda et al., 1997; Matsumoto et al., 2015). In accordance with these data, we found that strong induction of FasL-mediated apoptosis required a contact-dependent mechanism; while supernatants had less ability to kill HepG2-NTCP cells, and there was no significant FasL dependence. When put into context, CD8 T cells constituted the largest immune population in the liver and flare CD8 T cells were the largest cluster of CD8 T cells, suggesting they are sufficiently abundant to drive significant liver damage. In addition, our data suggest flare CD8 T cells can also mediate serial killing of hepatoma cells. We could induce FasL expression on 12.9% of CXCR6(+) CD8 T cells, which led to killing 50% of hepatoma cells, indicating each cell has the potential to kill multiple (in our model: 4, over 24 h) hepatocytes.

It is noteworthy that by week 12 of therapy, the majority of ALT decline had occurred, but we observed only modest changes in gene expression. Flare CD8 T cells were the only CD8 T population with a substantial number of downregulated genes during this window, suggesting it was primarily responsible for liver damage at baseline. However, even in flare CD8 T cells, FasL expression had only decreased 1.2-fold by week 12, with distinctly more changes in gene expression by week 24. We interpret this discrepancy as the activation of intrahepatic immunity lagging behind the clinical marker of liver damage, ALT decline. Consistent with the discordant kinetics in TAF-treated HBV patients, acute HCV infection in chimpanzees found upregulation of key inflammatory genes weeks to months before ALT elevation, whereas hepatocyte necrosis coincided with ALT elevation (Choi et al., 2016). This indicates that hepatocyte killing significantly lagged behind intrahepatic immune activation. Our data suggest prolonged intrahepatic immune activation as ALT approached normal values. This is important because ALT is not an ideal marker of intrahepatic immune activity (Cheong et al., 2011) and such a delay could have additional implications for using ALT as a marker when administering immunotherapies.

FNAs have been used extensively to sample tumors, for example in the kidneys (Turner et al., 2020; Eszlinger et al., 2017; Ono et al., 2020; Franzén et al., 2019; Su et al., 2021; Eikrem et al., 2018), but have recently gained traction for the study of liver disease due to their better tolerability for patients compared to liver biopsies. A finer gauge needle is used for liver FNAs than what is used for core liver biopsies, which reduces discomfort and allows for frequent sampling. The trade-off is the loss of tissue architecture, lack of high-quality hepatocytes and some blood contamination (Genshaft et al., 2021). However, an advantage demonstrated here is that liver FNAs can consistently capture the immunodiversity of the liver within patients across longitudinal time points. This is critical when investigating liver pathogenesis and will be important to understanding mechanisms of action of novel therapeutic agents that are currently in development to functionally cure the 290 million individuals living with chronic HBV infection (Gill et al., 2018; Gehring and Protzer, 2019). Another important advance is that cells collected from FNA passes can be cryopreserved similar to standard protocols for PBMC. Despite freezing <100,000 cells/vial, we could recover significant numbers of viable cells to validate longitudinal scRNAseq data by flow cytometry. This opens the window to analyze single cell data and return to cryopreserved samples to validate observations without having to enroll new patients for liver FNAs.

Our study combined longitudinal liver FNA sampling with state-of-the-art technology to investigate immunological mechanisms of liver pathogenesis in an investigator-initiated clinical trial. Our approach addressed key questions, so far only investigated in animal models, to define innate triggers that cause bystander activation of a defined CD8 T population capable of TCR-independent hepatocyte killing. Given that liver damage is the primary driver of disease progression, identifying sources of liver damage provides important knowledge to understand pathogenesis in the natural history of CHB, and mechanisms of therapeutic interventions. In conclusion, we demonstrate the value of combining longitudinal liver sampling, scRNAseq data analysis and validation using cryopreserved samples, which can serve as an example for future clinical trials.

## Methods

### Lead contact

Further information and requests for resources and reagents should be directed to and will be fulfilled by Adam Gehring.

### Materials and availability

Clinical samples cannot be made available due to limited amounts of CHB patients’ and IHMC donors’ material.

### Data and code availability

De-identified patient data will be shared by the lead contact upon request. Single-cell RNA sequencing data and original code will be shared by the lead contact upon request. Any additional information required to reanalyze the data reported in this paper is available from the lead contact upon request.

### Experimental models and patient details

#### Chronic hepatitis B patients

15 patients were included in this study to analyze blood, PBMC, and liver FNAs. Inclusion criteria were chronic hepatitis B (HBsAg(+) ≥ 6 months); age >18 years; high-normal or elevated ALT levels, defined as >19 IU/l for females and >30 IU/l for males (with ULN defined as >25 IU/l for females and >35 IU/l for males); HBV DNA >10000 IU/ml for HBeAg(+) and >1000 IU/ml for HBeAg(-) patients; adequate contraception. An overview of baseline characteristic is given in suppl. table 1. At baseline, all included patients had elevated ALT levels above the ULN. Exclusion criteria were antiviral or IFN treatment in the previous 6 months; immunosuppressive treatment in the previous 6 months; treatment with an investigational drug in the previous 3 months; history of decompensated liver cirrhosis; liver transplantation; co-infection with HCV, HDV, or HIV; other significant liver disease (such as alcoholic or drug-related liver disease, autoimmune hepatitis, hemochromatosis, Wilson’s disease or α1 antitrypsin deficiency); estimated glomerular filtration <50 ml/min/1.73m^2^ or significant renal disease; α-fetoprotein >50 ng/ml; pregnancy or breast feeding; other significant medical illness that might interfere with the study (e.g. immunodeficiency syndromes or malignancies); substance abuse.

Of the 5 patients analyzed by scRNAseq, 4 were male and one was female. Of the 4 patients analyzed for verification of the transcriptomic data on the protein level, 2 were male and 2 were female. Due to low numbers, no analysis of sex influence was included.

The study was approved by the University Health Network Research Ethics Board (CAPCR ID: 18-5748) and informed consent was obtained from all subjects. The study number is CO-US-320-4667.

#### Human donors of intrahepatic mononuclear cells (IHMCs)

6 healthy living liver donors were included to obtain IHMCs from liver perfusion solutions during the transplantation. Mean age at the time of donation was 39.8 years (range 30-53 years). 1 patient was male and 5 patients were female. Due to low numbers, no analysis of sex influence was included.

The study was approved by the University Health Network Research Ethics Board (CAPCR ID: 14-7425). Informed consent was obtained from all subjects.

#### HepG2-NTCP cells

The human hepatoma cell line HepG2-NTCP was originally obtained from a Caucasian American male adolescent (Knowles et al., 1980) and modified to overexpress the HBV entry receptor sodium taurocholate co-transporting polypeptide (NTCP) (Ni et al., 2014). HepG2-NTCP cells were kindly provided by Stephan Urban, Heidelberg. Cells were maintained in Dulbecco’s Modified Eagle Medium (DMEM) (Gibco), supplemented with 10% fetal bovine serum (FBS), 20 mM HEPES buffer, 100 U/ml Penicillin, 100 µg/ml Streptomycin, 10 ng/ml Plasmocin, 2% minimum essential medium (MEM) amino acids (Gibco), 1% MEM non-essential amino acids (Gibco), 1 mM Glutamax (Gibco), and 1 mM sodium pyruvate. When co-cultured with IHMC-derived cells, AIM-V medium (Gibco) + 100 µg/ml primocin + 2% human AB serum was used. All cells were grown and cultivated in an incubator holding a temperature of 37°C, a humidity of 95% and a CO2 fraction of 5%. HepG2-NTCP cells were passaged regularly, but no more than 12 times.

### Method details

#### Study design

This was an investigator-initiated, open-label phase 4 study at the Toronto Centre for Liver Disease, Canada. Patients started therapy with 25 mg/d TAF for the entire study duration of 48 weeks and were offered to continue therapy after the end of the study. Blood and FNA samples were collected at baseline, week 12 and week 24. Additional blood samples were collected at week 36 and 48.

#### Analysis of blood markers of HBV infection

ALT in patients’ blood was quantified using Advia (Siemens). HBV DNA was analyzed with AmpliPrep Taqman (Roche). HBsAg and HBeAg were measured by Architect assay (Abbott). These measurements were done by the Laboratory Medicine Program of Toronto General Hospital/University Health Network.

#### PBMC isolation

Blood was collected from CHB patients at the time of FNA collection, or from living liver donors just before transplantation. Density centrifugation was used to remove red blood cells: total blood was diluted in a 1:2 ratio with 2% Knockout Serum Replacement (Gibco) before layering on Lymphoprep solution (StemCell) in SepMate-50 tubes (StemCell). The supernatants containing PBMC were transferred and cells were washed twice with PBS+ 2% Knockout Serum Replacement and counted. Cells were cryopreserved in Knockout Serum Replacement + 10% dimethyl sulfoxide (DMSO).

#### Liver FNA collection

Liver FNAs were collected by a specialist. A suitable location was determined using ultrasound and anesthetized with lidocaine. 25-gauge needles were used for puncture and aspiration of cells into a 10 ml syringe that already contained 0.5 ml of RPMI medium (Gibco). After removal from the patient, the needle was flushed with additional 0.5 ml of medium to collect remaining cells. A total of four liver FNA passes was taken from each patient at each time point. Samples were maintained on ice and immediately processed as follows: the exact volume of each FNA pass was documented. Pictures were taken to document the color of each pass, indicating the blood content. In addition, a small fraction of each pass was used to collect optical density (OD) to obtain a quantitative measure of the blood content (Genshaft et al., 2021). For each analysis, we used the one or two passes with the lowest blood content from the respective timepoint and patient. Samples were counted and either cryopreserved in Knockout Serum Replacement (Gibco) + 10% DMSO for future analysis by flow cytometry or prepared for scRNAseq. For scRNAseq, red blood cells were removed by 5 min incubation with Red Blood Cell Lysis Buffer (BioLegend) and subsequent 10x dilution with PBS. Cells were washed twice, counted, and 20,000 cells per sample resuspended in 50 µl PBS + 0.04% BSA were subjected to scRNAseq.

#### Single-cell RNA sequencing on the 10x Genomics platform

Sample preparation: samples were prepared as outlined by the 10x Genomics Single Cell 5’ Reagent Kit user guide. Briefly, maximum volume was loaded to target capturing of max. 3,000 cells. After droplet generation, samples were transferred onto a pre-chilled 96-well plate, heat sealed and cDNA was generated overnight. The next day, cDNA was recovered using Recovery Agent provided by 10x Genomics, and subsequently purified using a Silane DynaBead (Thermo Fisher) mix as outlined by the user guide. Purified cDNA was amplified for 14 cycles before being purified again using SPRIselect beads (Beckman Coulter). Samples were run neat on a Bioanalyzer (Agilent Technologies) to determine cDNA concentrations. 5’ cDNA libraries were prepared as outlined by the 10x Genomics’ Single Cell 5’ Reagent Kit user guide, with modifications to the PCR cycles based on the calculated cDNA library input. To obtain TCR repertoire profiles from the same input samples, VDJ enrichment was performed with the Chromium Single Cell Human TCR Amplification Kit. Sequencing libraries were generated with unique sample indices for each sample and quantified.

Sequencing: The molarity of each library was calculated based on library size as measured by the Bioanalyzer (Agilent Technologies) and qPCR amplification data. Samples were pooled and adjusted to 10 nM, then diluted to 2 nM using elution buffer (Qiagen) + 0.1% Tween20. Each 2 nM pool was denatured by adding 0.1N NaOH at equal volumes for 5 minutes at room temperature. Library pools were further diluted to 20 pM using HT-1 (Illumina), and subsequently to a final loading concentration of 14 pM. 150 µl were loaded into each well of an 8-well strip tube and loaded onto a cBot (Illumina) for cluster generation. Samples were sequenced on the HiSeq 2500 (Illumina) system.

Unique molecular identifiers (UMIs) were generated using the CellRanger Pipeline version 3.1.0 (10x Genomics). Sequencing data were aligned to GRCh38-HBV reference genome.

#### Analysis of scRNAseq data

CellRanger-processed filtered feature matrices were analyzed using Seurat version 3.2.3 (Stuart et al., 2019). Data from individual samples was filtered to only preserve high-quality cells with >200 reads and <10% of mitochondrial DNA content; genes that appeared in less than 3 cells were filtered out. Data was normalized using Bioconductor’s scran with clusters (Lun et al., 2016), because normalization was shown to be the most influential step in scRNAseq analysis pipelines and scran was superior to other normalization methods (Vieth et al., 2019). Data from all samples was then integrated and scaled. Principal component analysis (PCA) dimensionality reduction was performed on the normalized gene expression counts. A nearest-neighbor graph was built with k.param set to 25 PCA dimensions as inputs. The neighborhood graph was used to calculate UMAP coordinates. Clusters were identified using Louvain algorithm with resolution set to 1. Cell types were annotated based on the canonical marker gene expression of each cluster. Whenever the canonical marker gene expression was ambiguous (this was the case for the cluster of proliferating cells), we used gene set enrichment analysis (GSEA) (Subramanian et al., 2005; Mootha et al., 2003) to identify pathways characterizing the cluster: gene rank lists were compiled in Seurat, GSEA was performed with GSEA version 4.0.3 (Broad Institute/UC San Diego), and enrichment maps were visualized using Cytoscape version 3.7.2.

Further analysis of scRNAseq data was done using R packages EnhancedVolcano version 1.11.3 (Blighe et al., 2021); scRepertoire version 1.3.1 (Borcherding et al., 2020); and NicheNet version 1.0.0 (Browaeys et al., 2020). Information on the respective vignettes is provided in the “additional resources” section.

#### Targeted gene expression scRNAseq

Targeted Gene Expression kits (Human Immunology Panel, 10x Genomics) were used for target enrichment and re-sequencing of libraries prepared for single-cell transcriptomic analysis. Experiments were performed according to manufacturer’s instructions. Briefly, sequencing libraries were quantified using an Agilent Bioanalyzer (High Sensitivity DNA Kit). If less than 300 ng DNA was available, the library was PCR-amplified using the Library Amplification Kit (10x Genomics). Libraries from a total cell count of 49,760 were pooled into three sets based on input cell counts: low (600–1400 cells/library), medium (1800–2500 cells/library) and high (4200–7700 cells/library). The dilution factors for pooling were calculated using the worksheet provided by the manufacturer. After adding Cot DNA and Universal Blockers, the library pools were dried in a vacuum centrifuge at 45°C (SpeedVac SPD210, Thermo Fisher). Target genes (1,056 immune-related genes) were enriched by hybridizing to gene-specific biotinylated baits bound to streptavidin beads. After washing capture beads, enriched libraries were PCR-amplified using the Library Amplification Kit with 10 total cycles and purified using SPRIselect reagent according to manufacturer’s protocol (Beckman Coulter). The enriched libraries were sequenced on NovaSeq platform (Illumina) with the sequencing depth of 10,000 read pairs per cell.

Sequencing data were aligned to GRCh38 reference genome and quantified using Cell Ranger version 6.0.1 (10x Genomics). Cells were filtered based on EmptyDrops method (Lun et al., 2019) implemented in Cell Ranger. After filtering, a total of 39,565 cells remained for downstream analysis. Seurat version 4.0 (Hao et al., 2021) was used to normalize cells using LogNormalize method (total UMI count for each cell was set to 10,000). PCA dimensionality reduction was performed on the normalized gene expression counts. A nearest-neighbor graph was built with k.param set to 20 and 30 PCA dimensions as inputs. The neighborhood graph was used to calculate UMAP coordinates. Clusters were identified using Louvain algorithm with resolution set to 0.8. Cell types were annotated based on the canonical marker gene expressions of each cluster.

#### Flow cytometric analysis of liver FNAs and matched PBMC

Preparation of liver FNAs was optimized to minimize cell loss. Longitudinal liver FNAs and matched PBMC from the same patients were thawed. Dead cells were stained with eFluor 520 (eBioscience) in PBS for 10 min. at room temperature. An equal volume of 2x concentrated extracellular antibodies in PBS was added for 30 min at 4°C: CD19_FITC; CD20_FITC; CD33_BB515; CD3_BV510; CD4_BUV395; CD8_APC-H7; γδTCR_BUV563; CD56_APC; CXCR6_BV421; CD27_PE-Cy7; CD38_BV786; 4-1BB_BV650; CD6_BUV737; FasL_BV605; PD-1_BB700; Lag3_APC-R700. Further information on antibodies used is listed in the key resources table. Cells were washed once with PBS + 0.5% bovine serum albumin (BSA) and then fixed and stored in PBS + 1% paraformaldehyde (PFA) for flow cytometry analysis. Analysis was done using an LSR Fortessa (Becton Dickinson) cytometer. Data was analyzed with FlowJo version 10.7.1.

#### Analysis of clusters defined by flow cytometry using flowCore and CATALYST

Flow cytometry data was further analyzed using R packages flowCore version 2.5.0 (Ellis et al., 2021) and CATALYST version 1.17.3 (Crowell et al., 2021). Single, live, CD3(+) lymphocytes were gated in FlowJo and fcs files were imported into R without truncation for further analysis. Data from FNA or PBMC samples from the same timepoint were clustered using a 10×10 grid and evaluation of 2 through 20 metaclusters. A delta area plot was used to determine the optimal number of clusters. UMAP dimensional reduction was performed on all cells.

#### Living liver donor IHMC collection

IHMC were isolated from living donor liver transplantation perfusions at Toronto General Hospital. Prior to transplantation, livers were perfused with 1-2 L of cold University of Wisconsin (UW) solution (ViaSpan) to remove immune cells from the sinusoids. Perfusates were collected and centrifuged for concentration in HBSS + 2 U/ml heparin, to a final volume of 50-100 ml. Cells were then subjected to density centrifugation by layering cells on Lymphoprep solution (StemCell) in SepMate-50 tubes (StemCell) to remove red blood cells. Supernatants containing IHMC were washed 2x with PBS + 2% Knockout Serum Replacement (Gibco). Cells were counted and cryopreserved in Knockout Serum Replacement + 10% DMSO.

#### Cytokine stimulation of IHMC and flow cytometry analysis

IHMC were maintained in AIM-V medium + 100 µg/ml primocin + 2% human AB serum. After thawing, they were stained with CXCR6_BV421, 2.5:100, diluted in medium, for 30 min at 4°C. They were then adjusted to a concentration of 1.5 million cells/ml for cultivation and cytokine treatment. Indicated cytokines were used at the following concentrations: IL-2 (GoldBio) at 100 IU/ml; IL-12p70 (BioLegend) at 25 ng/ml. Additional cytokines were used in exploratory experiments: IL-4 25 ng/ml; IL-10 25 ng/ml; IL-15 25 ng/ml; IL-21 25 ng/ml; IL-27 25 ng/ml; TGFβ 10 ng/ml; TNFα 100 ng/ml; IFNα 100 IU/ml; IFNγ 100 IU/ml. Further information on the cytokines used is listed in the key resources table. IHMC were treated for 24 h before supernatants were collected for further HepG2-NTCP treatment. Cells were collected for flow cytometry analysis, or for sorting and subsequent co-cultivation with HepG2-NTCP cells.

For flow cytometry analysis, eFluor 506 (eBioscience) in PBS was used for staining of dead cells for 10 min at room temperature. Cells were then incubated with extracellular antibodies for 30 min at 4°C: CD8_APC-H7; CXCR6_BV421; and FasL_BV605 (all antibodies were diluted in staining buffer: PBS + 1% BSA + 0.1% sodium azide). We performed CXCR6 staining both before and after cultivation to capture as many CXCR6(+) cells as possible. Cells were permeabilized using Cytofix/Cytoperm (BD Biosciences) for 15 min at 4°C, followed by incubation with intracellular antibodies: IFNγ_APC; 2.5:100, and FasL_BV605, 2.5:100 (all antibodies diluted in PermWash buffer: PBS + 1% BSA + 0.1% sodium azide + 0.1% saponin). Suppliers of all antibodies are listed in the key resources table. Cells were washed once with staining buffer, fixed and stored in PBS + 1% PFA. Flow cytometry was performed using an LSR II OICR, BGRV (Becton Dickinson) cytometer.

For flow cytometry analysis using the multi-color panel with markers defining the flare CD8 T population, IHMC were thawed, stained with CXCR6_BV421, and treated for 24 h as described above. Dead cells were then stained with eFluor 520 (eBioscience), 1:1000 in PBS, for 10 min at room temperature. Subsequently, cells were stained with extracellular antibodies listed above (section “Flow cytometric analysis of liver FNAs and matched PBMC”) for 30 min at 4°C. Cells were washed once with PBS + 0.5% BSA and then fixed and stored in PBS + 1% PFA for flow cytometry analysis. The LSR Fortessa (Becton Dickinson) cytometer was used. Data was analyzed with FlowJo version 10.7.1, and with flowCore version 2.5.0 and CATALYST version 1.17.3 as described above (section “Analysis of clusters defined by flow cytometry using flowCore and CATALYST”).

#### Sorting of IHMC to select CD8(+) CXCR6(+) cells

After cytokine stimulation for 24 h, IHMC were sorted prior to co-cultivation with HepG2-NTCP cells. IHMC were kept on ice during preparation and before/after sorting unless indicated otherwise. They were stained with viability dye eFluor520 1:1000 in PBS for 10 min at room temperature. Staining was performed by directly adding antibodies CD8_APC, final concentration 0.2:100 and CXCR6_BV421, final concentration 2.5:100 for 30 min at 4°C. They were washed once with PBS and resuspended in MACS buffer (PBS + 0.5% BSA + 2 mM EDTA) in tubes that had previously been blocked with PBS + 2% BSA. Directly prior to sorting, cells were filtered through a 70 µM sterile filter. We used a Sony SH800, BRV cell sorter to select live, CD8(+) CXCR6(+) cells. Sorted cells were counted and re-suspended in AIM-V + 100 µg/ml primocin + 2% human AB serum. Depending on the setup, they were not or were pre-incubated with 10 µg/ml neutralizing anti-FasL antibody (BioLegend) for 30 min at 37°C. Cells without and with anti-FasL were then added to the HepG2-NTCP cells for 24 h of co-cultivation.

#### HepG2-NTCP killing assay

HepG2-NTCP cells were seeded at 50,000 cells/well in a 96-well plate one day prior to co-culture with IHMC-derived CD8 T cells. Sorted CXCR6(+) CD8 T cells were then added to the HepG2-NTCP cells in a 1:1 ratio and cultured in AIM-V + 100 µg/ml primocin + 2% human AB serum for 24 h. In parallel, HepG2-NTCP cells were cultured in the IHMC-derived supernatants (50 µl/well) for 24 h. Adherent HepG2-NTCP cells were detached from the cell culture plates by incubation with 0.5 mM EDTA (Invitrogen) for 10 min at 37°C. Dead cells were stained using eFluor506 (eBioscience) 1:500 in PBS (Gibco) for 10 min at room temperature. Cells were then fixed with PBS + 1% PFA (Sigma-Aldrich) in PBS for 15 min at 4°C. To permeabilize cellular membranes, including mitochondrial and nuclear membranes, cells were incubated with BD Phosflow PermBuffer III (BD Biosciences), containing 87.86% methanol, for 30 min on ice. They were washed with PBS + 0.5% BSA before incubation with the cleaved caspase 3_AF647 antibody, 0.5/100 diluted in PBS + 0.5% BSA for 30 min at 4°C. Cells were washed again and stored in PBS + 1% PFA in the dark before flow cytometry analysis. Cells were analyzed using an LSR II OICR, BGRV (Becton Dickinson) cytometer and FlowJo version 10.7.1.

## Statistical analysis

Statistical analysis was done using GraphPad Prism 8 and R version 4.1.0. Statistical tests used and numbers of replicates are indicated in the respective results sections and figure legends.

## Supporting information

Suppl. methods_key resources table

## Data Availability

All data produced in the present study are available upon reasonable request to the authors.

## Additional resources

Seurat vignette: https://satijalab.org/seurat/articles/get_started.html

scran vignette: https://rdrr.io/bioc/scran/f/vignettes/scran.Rmd

scRepertoire vignette: https://ncborcherding.github.io/vignettes/vignette.html

EnhancedVolcano vignette: https://bioconductor.org/packages/release/bioc/vignettes/EnhancedVolcano/inst/doc/En hancedVolcano.html

NicheNet vignette: https://rdrr.io/github/saeyslab/nichenetr/f/vignettes/seurat_wrapper.md

flowCore vignette: https://github.com/RGLab/flowCore/blob/master/vignettes/HowTo-flowCore.Rnw

CATALYST vignette: https://bioconductor.org/packages/release/bioc/vignettes/CATALYST/inst/doc/differential.html

## Notes

### Competing Interest Statement

S.N., A.K., J.D.S.V., A.M., A.P., C.H., and I.M. have no competing interests to declare.
D.M.: employed by Fluidigm Inc.
S.C.K.: employed by and stockholder of Gilead Sciences Inc.
J.J.F.: research funding by Abbvie, Arbutus Biopharma, Gilead Sciences Inc., Janssen Pharmaceuticals, Eiger Biopharmaceuticals, and Enanta Pharmaceuticals; consulting/scientific advising for Abbvie, Arbutus Biopharma, Gilead Sciences Inc., and GlaxoSmithKline.
S.F.: research funding by Gilead Sciences Inc.; speakers bureau/honoraria: Gilead Sciences Inc., and Abbvie; consulting/scientific advising for Gilead Sciences Inc., Abbvie, Janssen Pharmaceuticals, Assembly Biosciences.
D.C.: formerly employed by Gilead Sciences Inc. and currently employed by Bristol Myers Squibb.
J.J.W.: employed by and stockholder of Gilead Sciences Inc.
A.G.: formerly employed by and stockholder of Gilead Sciences Inc.
H.L.A.J.: research funding by Abbvie, Gilead Sciences Inc., GlaxoSmithKline, Janssen Pharmaceuticals, Roche, Vir Biotechnology; consulting/scientific advising for ALIGOS Therapeutics, Antios Therapeutics, Arbutus Biopharma, Eiger Biopharmaceuticals, Gilead Sciences Inc., GlaxoSmithKline, Janssen Pharmaceuticals, Merck, Roche, VBI Vaccines Inc., Vir Biotechnology, and Viroclinics Biosciences.
A.J.G.: research funding by Janssen Pharmaceuticals, GlaxoSmithKline, and Gilead Sciences Inc.; consulting/scientific advising: Janssen Pharmaceuticals, Roche, GlaxoSmithKline, Vir Biotechnology, Finch Therapeutics, SQZ Biotech.

### Clinical Trial

CO-US-320-4667

### Funding Statement

This is an investigator-intitated study funded by Gilead Sciences Inc.

### Author Declarations

The University Health Network Research Ethics Board (University Health Network, Toronto, Canada) gave ethical approval for this work.

