## Supplementary material for "Pathogenic CD8 T cells defined by longitudinal liver sampling in chronic hepatitis B patients starting antiviral therapy": Suppl. methods_key resources table

| REAGENT or RESOURCE | SOURCE | IDENTIFIER |
|---|---|---|
| **Antibodies** | | |
| CD3_BV510, mouse anti-human | BD Biosciences | Cat#563109<br>Clone UCHT1 |
| CD4_BUV395, mouse anti-human | BD Biosciences | Cat#563550<br>Clone SK3 |
| CD6_BUV737, mouse anti-human | BD Biosciences | Cat#749476<br>Clone M-T605 |
| CD8_APC, mouse anti-human | BD Biosciences | Cat#561421<br>Clone RPA-T8 |
| CD8_APC-H7, mouse anti-human | BD Biosciences | Cat#560179<br>Clone SK1 |
| CD19_FITC, mouse anti-human | BioLegend | Cat#302206<br>Clone HIB19 |
| CD20_FITC, mouse anti-human | BioLegend | Cat#302304<br>Clone 2H7 |
| CD27_PE-Cy7, mouse anti-human | BD Biosciences | Cat#560609<br>Clone M-T271 |
| CD33_BB515, mouse anti-human | BD Biosciences | Cat#564588<br>Clone WM53 |
| CD38_BV786, mouse anti-human | BD Biosciences | Cat#563964<br>Clone HIT2 |
| CD56_APC, mouse anti-human | BD Biosciences | Cat#555518<br>Clone B159 |
| 4-1BB (CD137)_BV650, mouse anti-human | BD Biosciences | Cat#564092<br>Clone 4B4-1 |
| Fas ligand (CD178)_BV605, mouse anti-human | BD Biosciences | Cat#744099<br>Clone NOK-1 |
| CXCR6 (CD186)_BV421, mouse anti-human | BD Biosciences | Cat#566007<br>Clone 13B 1E5 |
| Lag3 (CD223)_APC-R700, mouse anti-human | BD Biosciences | Cat#565774<br>Clone T47-530 |
| PD-1 (CD279)_BB700, mouse anti-human | BD Biosciences | Cat#566460<br>Clone EH12.1 |
| gdTCR_BUV563, mouse anti-human | BD Biosciences | Cat#748534<br>Clone 11F2 |
| IFNg_APC, mouse anti-human | Biolegend | Cat#506510<br>Clone B27 |
| Cleaved Caspase-3_AF647, rabbit anti-human | Cell Signaling | Cat#9602S<br>Lot 9 |
| viability dye eFluor506 | eBiosciences | Cat#65-0866-14 |
| viability dye eFluor520 | eBiosciences | Cat#65-0867-14 |
| Fas ligand (CD178) Ultra-LEAF Purified, mouse anti-human | BioLegend | Cat#306415<br>Clone NOK-1 |
| **Chemicals, Peptides, and Recombinant Proteins** | | |
| IL-2 | Goldbio | Cat#1110-02-50 |
| IL-4 | Goldbio | Cat#1110-04-5 |

| | | |
|---|---|---|
| IL-10 | Goldbio | Cat#<br>1110-10-2 |
| IL-12p70 | BioLegend | Cat#573002 |
| IL-15 | Goldbio | Cat#<br>1110-15-10 |
| IL-21 | Goldbio | Cat#<br>1110-21-10 |
| IL-27 | Biolegend | Cat#<br>589202 |
| TGFβ | StemCell | Cat#78067.1 |
| TNFα | Goldbio | Cat#1130-01-10 |
| IFNα | Goldbio | Cat#1160-03-20 |
| IFNγ | Goldbio | Cat#1160-06-20 |
| **Experimental Models: Cell Lines** | | |
| HepG2-NTCP | kindly provided by Stephan Urban, Heidelberg | |
| **Software and Algorithms** | | |
| R versions 3.6.3 – 4.1.0 | The R Project | https://www.r-project.org/ |
| Seurat versions 3.2.3 and 4.0 | Satija lab | https://satijalab.org/seurat/index.html<br>Stuart et al., 2019<br>Hao et al., 2021 |
| scran | Bioconductor | https://bioconductor.org/packages/release/bioc/html/scran.html<br>Lun et al, 2016 |
| EnhancedVolcano version 1.11.3 | Kevin Blighe | https://github.com/kevinblighe/EnhancedVolcano<br>Blighe et al., 2021 |
| scRepertoire | Nick Borcherding | https://github.com/ncborcherding/scRepertoire<br>Borcherding et al., 2020 |
| NicheNet | Robin Browaeys | https://github.com/saeyslab/nichenetr<br>Browaeys et al., 2020 |
| flowCore version 2.5.0 | Bioconductor | https://bioconductor.org/packages/release/bioc/html/flowCore.html<br>Ellis et al., 2021 |
| CATALYST version 1.17.3 | Bioconductor | https://bioconductor.org/packages/release/bioc/html/CATALYST.html<br>Crowell et al., 2021 |
| GraphPad Prism 8 | GraphPad | |
| FlowJo version 10.7.1 | FlowJo, LLC | |
