## Supplementary material for "Pathogenic CD8 T cells defined by longitudinal liver sampling in chronic hepatitis B patients starting antiviral therapy": Suppl. table and figures

**Supplemental information**

**Suppl. table 1. Characteristics of included patients with chronic hepatitis B.**

| | Mean | Range |
|---|---|---|
| Age [years] | 44.3 | 29–64 |
| Male sex [% of all patients] | 60% | |
| ALT at baseline [× ULN] | 4.4 | 1.1–21.8 |
| HBV DNA at screening [IU/ml] | $4.84 \times 10^{6}$ | $1.89 \times 10^{4}$–$7.97 \times 10^{7}$ |
| Baseline HBeAg(+) [% of all patients] | 40% | |

**Suppl. Figure 1. Populations obtained from scRNAseq were equally distributed among all patients.** (A) UMAP plots, corresponding to the one in Figure 2A, for each individual patient. (B) Distribution of clusters among all cells, by patient. There was no statistical difference in the distribution of clusters across patients.

**Suppl. Figure 2. Gating strategy used for clustering of flow cytometry data.** Multi-color flow cytometry of one representative FNA baseline sample.

**Suppl. Figure 3. Tissue residency characteristics of intrahepatic lymphocytes, compared to PBMC lymphocytes.** (A) Flow cytometry of FNA and matched PBMC of 4 CHB patients before and during TAF therapy; and of 5 living liver donor IHMC. CXCR6 as a marker of tissue residency was quantified in CD8 T cells. (B) Quantification of CD4 and CD8 T cells in PBMC and matched intrahepatic cells from living liver donors. (C) CD69 and CD103 in CD4 and CD8 T cells derived from PBMC, or matched IHMC.

**Suppl. Figure 4. The combination of IL-2 + IL-12 is most effective in inducting key markers of flare CD8 T cells in healthy donor IHMC.** Cytokines that had been *in silico* predicted to upregulate IFNγ and Fas ligand, key markers of flare CD8 T cells in CHB patients at baseline, were assessed with respect to their potential to upregulate those markers in healthy donor IHMC. IHMC were treated for 24 h with the indicated cytokines before IFNγ (A-B) and Fas ligand (C-D) were quantified in CXCR6(+) CD8 T cells by flow cytometry. (A, C) individual predicted markers, (B, D) selected combinations of 2 and 3 different cytokines.

**Suppl. Figure 5. Flare CD8 T markers in healthy donor-derived IHMC induced by IL-2 and IL-12.** Multi-color flow cytometry of one representative donor. Gating: Lymphocytes → single cells → live cells → CD3(+) → CD8(+) and CXCR6(+) → phenotypic markers that define the flare CD8 T population.

### Suppl. figure 1

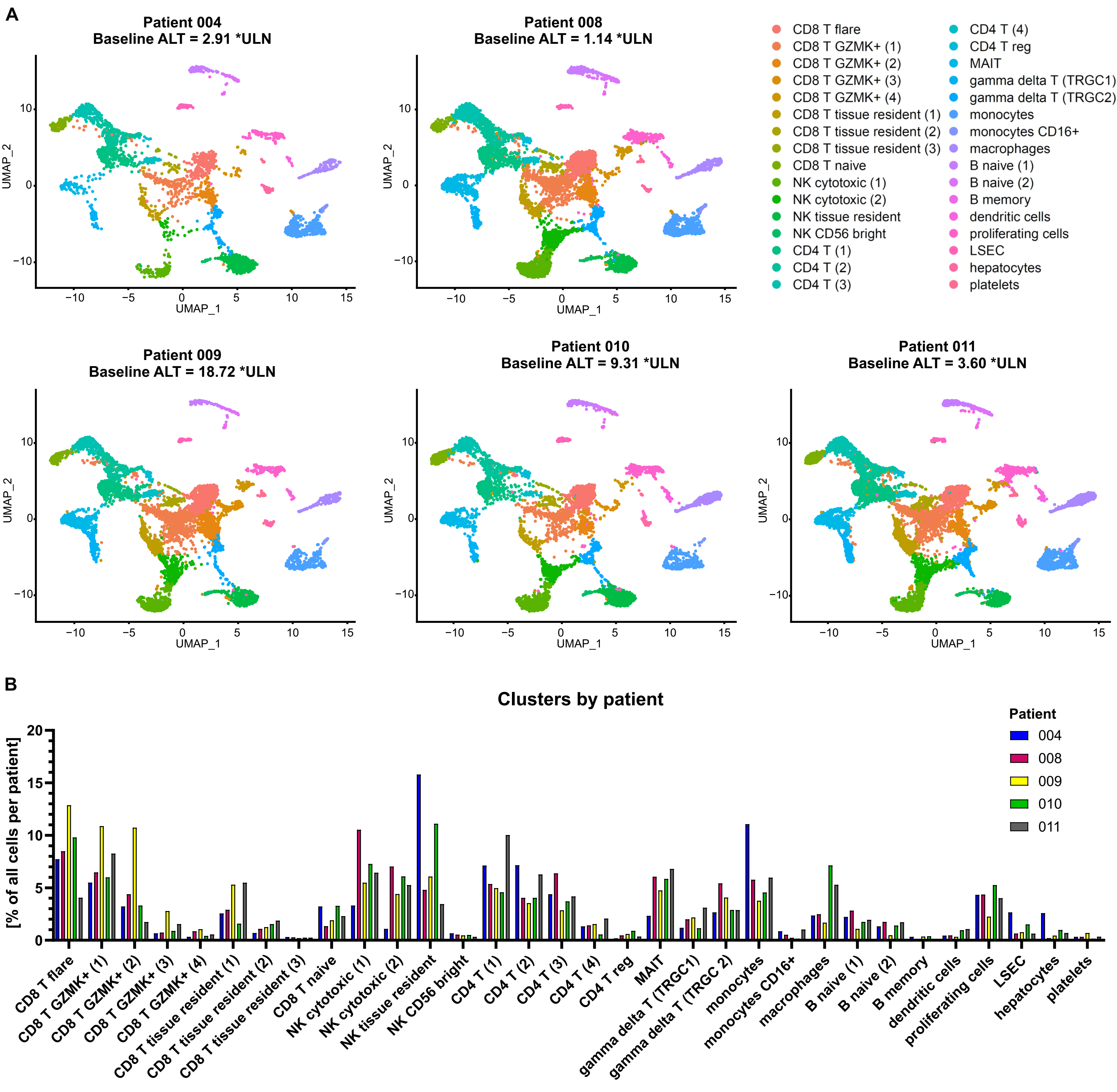

### Suppl. figure 2

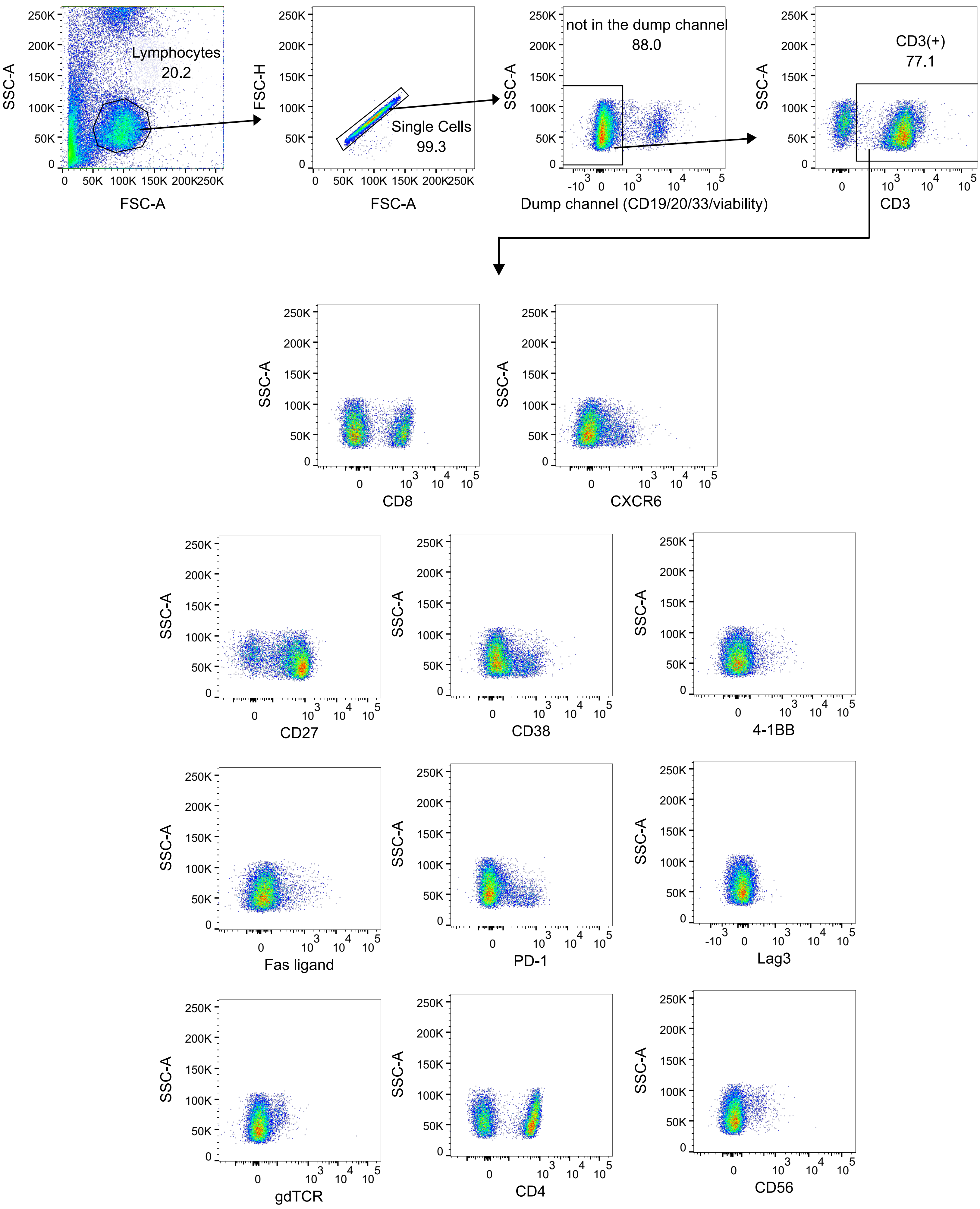

**Suppl. figure 3**

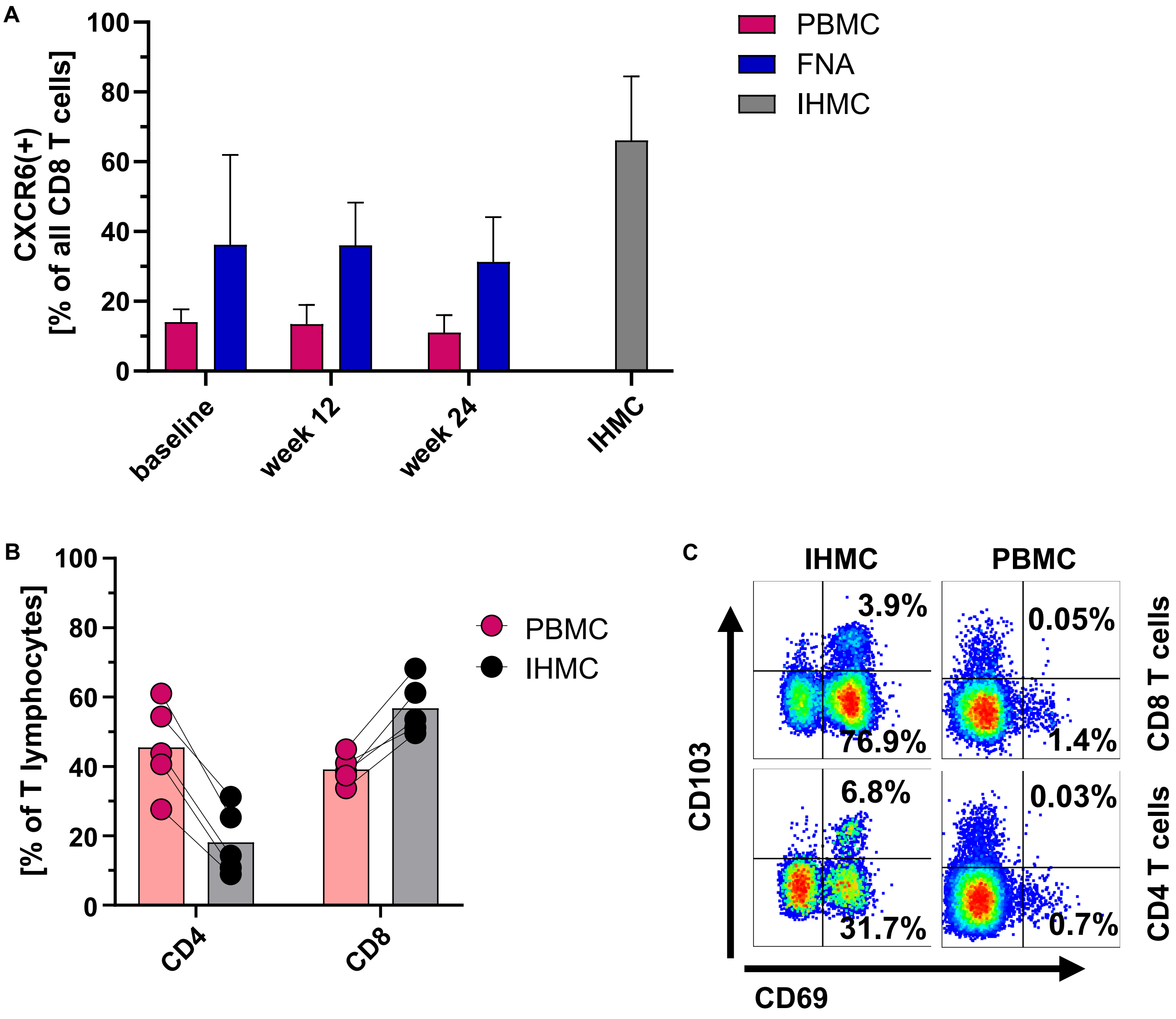

#### Suppl. figure 4

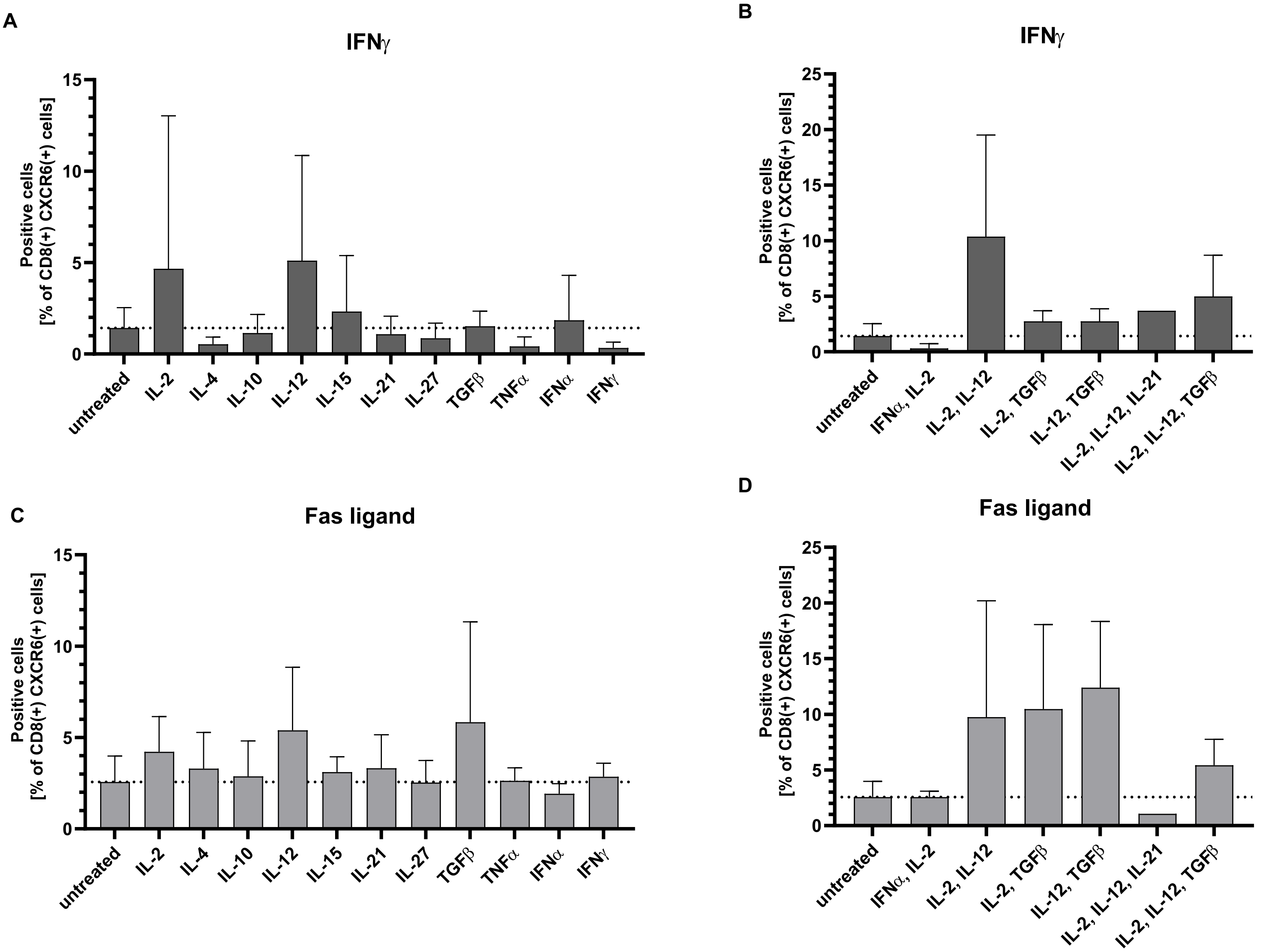

### Suppl. figure 5

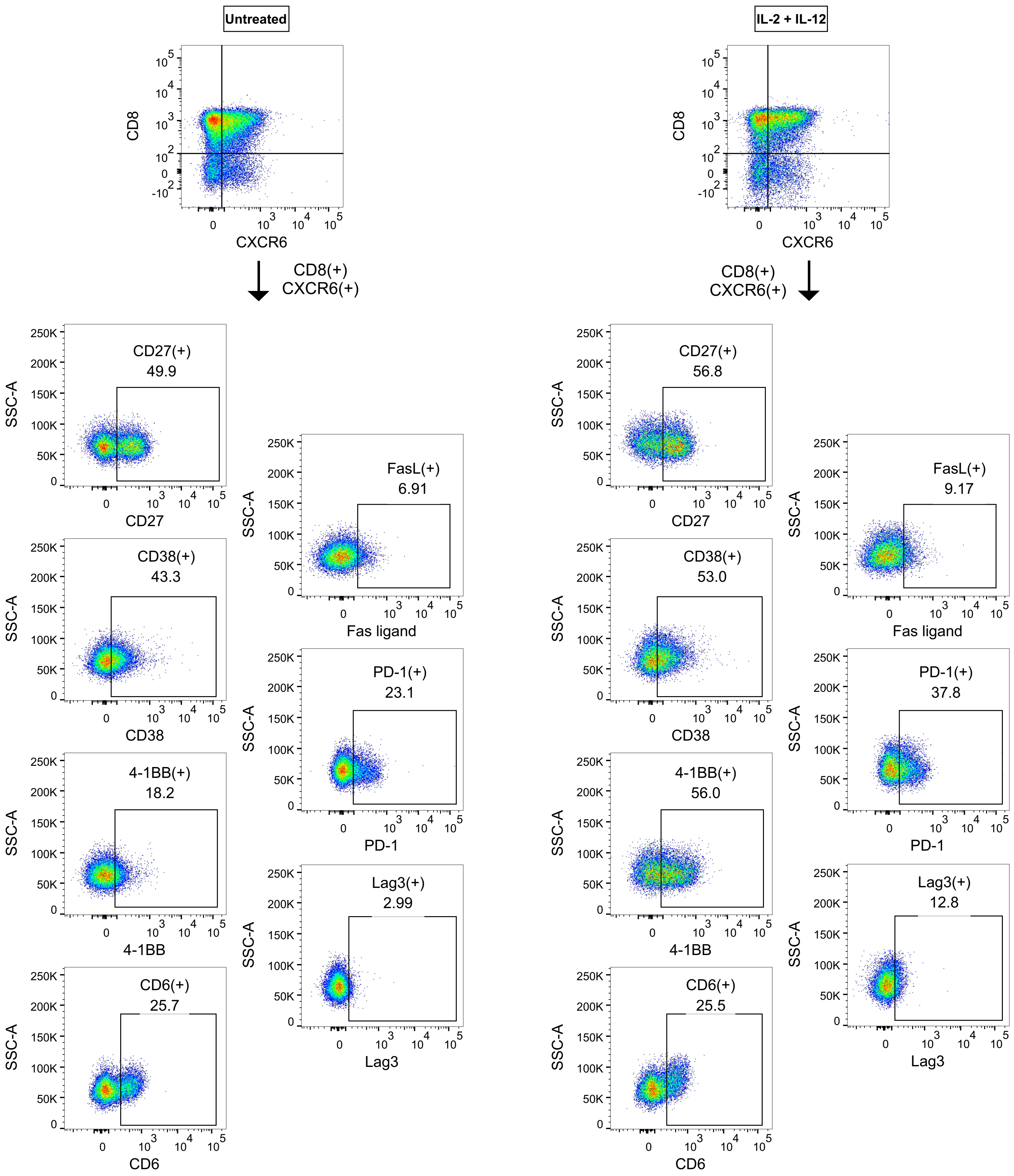
